# Human blood vessel organoids reveal a critical role for IGFBP7 in Diabetic Microangiopathy

**DOI:** 10.64898/2026.07.26.26358752

**Authors:** Aman Pooni, Konstantinos Theofilatos, Ludovic Boas, Thakorn Pruktanakul, Sharmin Alhaque, Sara G Romeo, Ilaria Secco, Edoardo Schneider, Rafael Oexner, Robin Schmitt, Anna Zoccarato, Seong-Hun Park, Steven Lynham, Lingfang Zeng, Chan Ning Lee, Timothy L. Jackson, Bijan Modarai, Andriana Margariti, Mauro Giacca, Ajay M Shah, Anna Zampetaki

## Abstract

Microangiopathy is a key contributor to diabetic vascular complications. There is currently no effective treatment, and patients are at high risk of major adverse cardiovascular events. We have recently demonstrated that human blood vessel organoids (BVOs) capture key aspects of the microvasculature. Here, employing patient derived BVOs we find that diabetic media treated BVOs display features of diabetic clinical microvascular specimens. Proteomic profiling of diabetic BVOs and their secretome, revealed a robust upregulation of IGFBP7, a protein that was subsequently associated with microangiopathy in UK Biobank. Follow-up studies uncovered an instrumental role of IGFBP7 in the microvasculature and LGALS1 and LGALS3 as its downstream effectors. Underlying their importance in a clinical setting, in the UKB biobank, we observed a strong association of LGALS1, LGALS3 with diabetic nephropathy and retinopathy while in AoU cohort we identified genetic variants in the diabetic microangiopathy signature associated with diabetes diagnosis and cardiometabolic outcomes.

## Introduction

Diabetic microangiopathy is a major complication of diabetes mellitus. It can occur in multiple vascular beds (retina, kidneys, nerves and skin)^1^ and its progression rate differs from patient to patient. The duration and severity of hyperglycemia are thought to be important determinants of vascular injury, but the mechanisms of disease are not well understood^2^.

Microangiopathy is triggered by two early restructuring events. The disruption of endothelial cell (EC): pericyte (PC) interactions that leads to vessel regression^2^ and the alterations in extracellular matrix (ECM)^3^ that affect vessel contractility are the two critical changes in the microvasculature that can cause local ischemia^4^. These structural readouts that indicate functional changes can be quantified in human blood vessel organoids (BVOs)^5^. Derived from induced pluripotent stem (iPS) cells, BVOs can recapitulate the morphological and molecular features of human microvasculature forming capillaries with a lumen, endothelial lining, PC coverage and a basement membrane^6^. Amenable to genetic manipulation they are suitable for mechanistic studies into human pathophysiology, can accelerate drug development and facilitate personalised treatment approaches. Hence, structural assessment of BVOs can offer an innovative platform for functional characterization of the human microvasculature^6^.

Recently, we showed that human BVOs undergo prompt restructuring in response to metabolic changes^5^. Targeting of PFKFB3, an activator of glycolysis, led to BVO remodelling and vessel regression with reduced PC coverage. Their rapid response renders BVOs particularly attractive for mechanistic studies and the identification of potential targets for novel therapeutic approaches. Notably, the short timeframe of response is extremely useful and desirable for drug and small compound screening applications^5^.

Here, we use BVOs to establish the molecular signature of diabetic microangiopathy and uncover novel events that can be targeted pharmacologically to restore the function of the microvasculature. To this end, we used diabetic patient and healthy (non-diabetic) donor-derived BVOs and treatment with diabetic media (high glucose, low cytokines), to recapitulate the events of early microangiopathy. Proteomic analysis of BVOs and their secretome revealed a profound remodelling in diabetic ECM and identified an instrumental role of insulin-like growth factor binding protein-7 (IGFBP7) and CD93 in the vasculature. IGFBP7 is a secreted protein previously linked to senescence, cardiovascular ageing, cardiac remodelling and myocardial dysfunction^7^. CD93 is involved in angiogenesis and can act as a soluble growth factor and an adhesion molecule^8^. The expression of both effectors and their link to diabetic microangiopathy was interrogated in the UK Biobank, a large prospective depository that includes screening data from hundreds of people living with diabetes and microvascular complications. Further mechanistic studies using engineered IGFBP7 knockout BVOs elucidated the key regulators in this process and, for the first time, highlighted LGALS1 and LGALS3 as downstream effectors of IGFBP7 and key mediators of vascular regression in diabetic microangiopathy.

## Results

### BVOs as a model of early events in diabetic microangiopathy

To determine whether BVOs can recapitulate the early events in diabetic microangiopathy, namely PC dropout and basement membrane thickening, we generated BVOs from human KOLF2 iPS cells using the Wimmer et al (2019) protocol^6^ and performed a 7-day treatment with media supplemented with high glucose (20mM) and low cytokines (1ng/ml IL6 and TNFα). This media was termed ‘diabetic media’, its glucose levels model the grade 4 serum hyperglycemia in patients and the proinflammatory cytokines, the increased levels of IL6 and TNFα that are observed in the vitreous and plasma patients with diabetic retinopathy^6,9^. Media supplemented with mannitol only, as an osmotic control, was termed ‘normal media’. We selected to test the 7-day timepoint, as this is a relatively short treatment that would be ideal for a high throughput drug screening pipeline.

Immunofluorescence staining demonstrated a dense vascular network consisting of CD31^+^ EC networks with PDGFR-β^+^ PCs attached to the microvessels (Fig. 1a). Flow cytometry analysis of BVOs digests further confirmed the presence of CD31^+^ ECs, mural cells as PDGFRβ^+^ cells, mesenchymal stem-like cells as CD90^+^CD73^+^CD44^+^ cells and haematopoietic cells as CD45^+^ (Fig. 1b). Interestingly, no statistically significant differences in the cellular composition were detected. Notably, in diabetic media treated BVOs a small portion of cells (∼5%) remained unlabelled. Diabetic media treatment did not affect the BVO diameter (Fig. 1c), vessel density or length (Fig. 1d, e), but led to a pronounced PC dropout in diabetic BVOs (Fig. 1f). Furthermore, ColIV, a key component of the basement membrane, was increased in the diabetic BVOs (Fig. 1g, h). Therefore, the 7-day diabetic media treatment of BVOs is a model that can capture the key early events in diabetic microangiopathy, namely PC dropout and ECM deposition.

**Figure 1.**
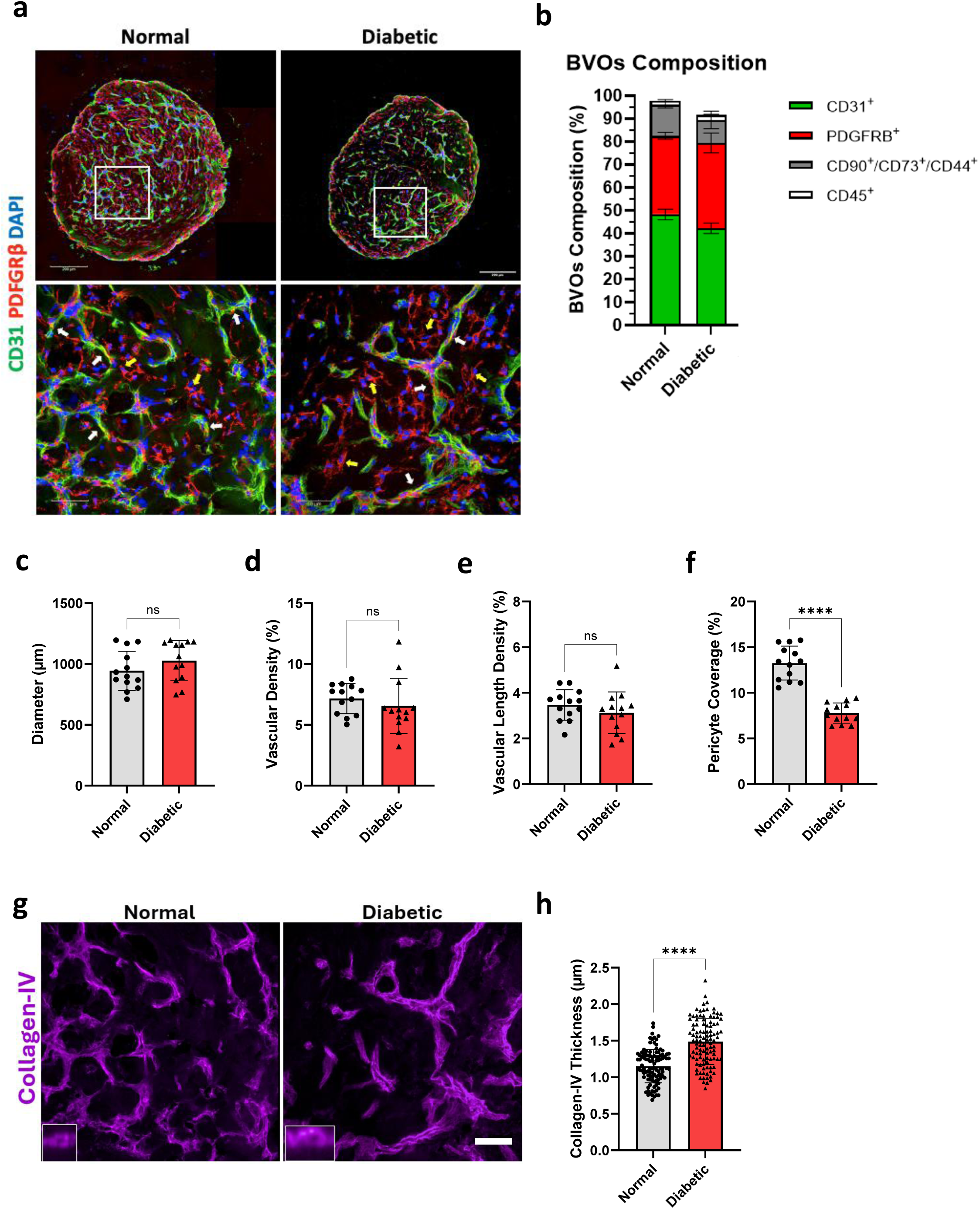
Diabetic media treated BVOs can recapitulate PC dropout and ECM deposition, two early events in diabetic microangiopathy. (a) Immunofluorescence confocal imaging of BVOs showing PC coverage (PDGFRβ^+^, red) of CD31^+^ ECs (green) following culture in normal and diabetic media for 7 days. The KOLF2 iPS line was used to generate these BVOs. White arrows indicate pericytes attached to a microvessel and yellow arrows indicate extravascular mural cells. (b) FACS was used to determine the different cell populations in BVOs. ECs were defined as CD31^+^, PCs as PDGFRβ^+^, mesenchymal stem-like cells as CD90^+^CD73^+^CD44^+^ and haematopoietic cells as CD45^+^. Cells from a total of 12-20 BVOs were dissociated and pooled together for this analysis from 3 independent preparations. (c) Quantification of BVO diameter from n=7 BVOs per group. One-two sections per BVO were assessed. (d) Quantification of vascular density and (e) vascular length density from n=7 BVOs per group, in three areas per 10x image. One-two sections per BVO were assessed. (f) Percentage of PC coverage from n=7 BVOs per group. One-two sections per BVO were assessed. (g) Immunofluorescence confocal imaging of BVO sections showing collagen-IV deposition (magenta). Insets show vessel cross sections. (h) Quantification of collagen-IV thickness measured by re-slicing z-stacks on the vertical axis where n=105 cross-sections from 7 BVOs per group. Data are presented as mean±SD. P-values were calculated using two-tailed Student’s t-test. (****p< 0.0001). Scale bars 200µm and 50µm.

### Generation of vascular cells from patient iPS

Patient-derived iPS cells are an attractive model of disease as they can reflect both the human heterogeneity and the epigenetic modifications triggered by the disease state^10^. Employing a stepwise differentiation of iPS cells to mesoderm and vascular lineages followed by MACS^11^ we isolated CD144^+^ iPS-derived endothelial cells (iPS-ECs) and CD144^-^/ PDGFRβ^+^ iPS-derived mural cells (iPS-MCs) (Fig. 2a). Similar to the KOLF2 reference line^5^, differentiation to vascular cells was efficient for patient lines (n=4) (Fig. 2b). For iPS-ECs, immunofluorescence staining confirmed successful differentiation, detecting the EC markers CD31, CD144 and tight junction protein 1 (ZO1) in the iPS-EC cells (Fig. 2c). ZO1 is a structural adaptor protein with a key role in tight junction function and complex formation of F-actin, transmembrane and cytosolic proteins^12^. Compared to healthy donor-derived iPS-ECs, diabetic donor cells displayed modifications in tight junction morphology. An increase in thick junctions, typically linked with increased permeability and a reduction in fingers junctions, which are associated with tip cells and migration^5,12^ (Fig. 2d, e) were observed. These findings in iPS-ECs cultured under baseline conditions imply the presence of metabolic memory in the diabetic donor derived cells.

**Figure 2.**
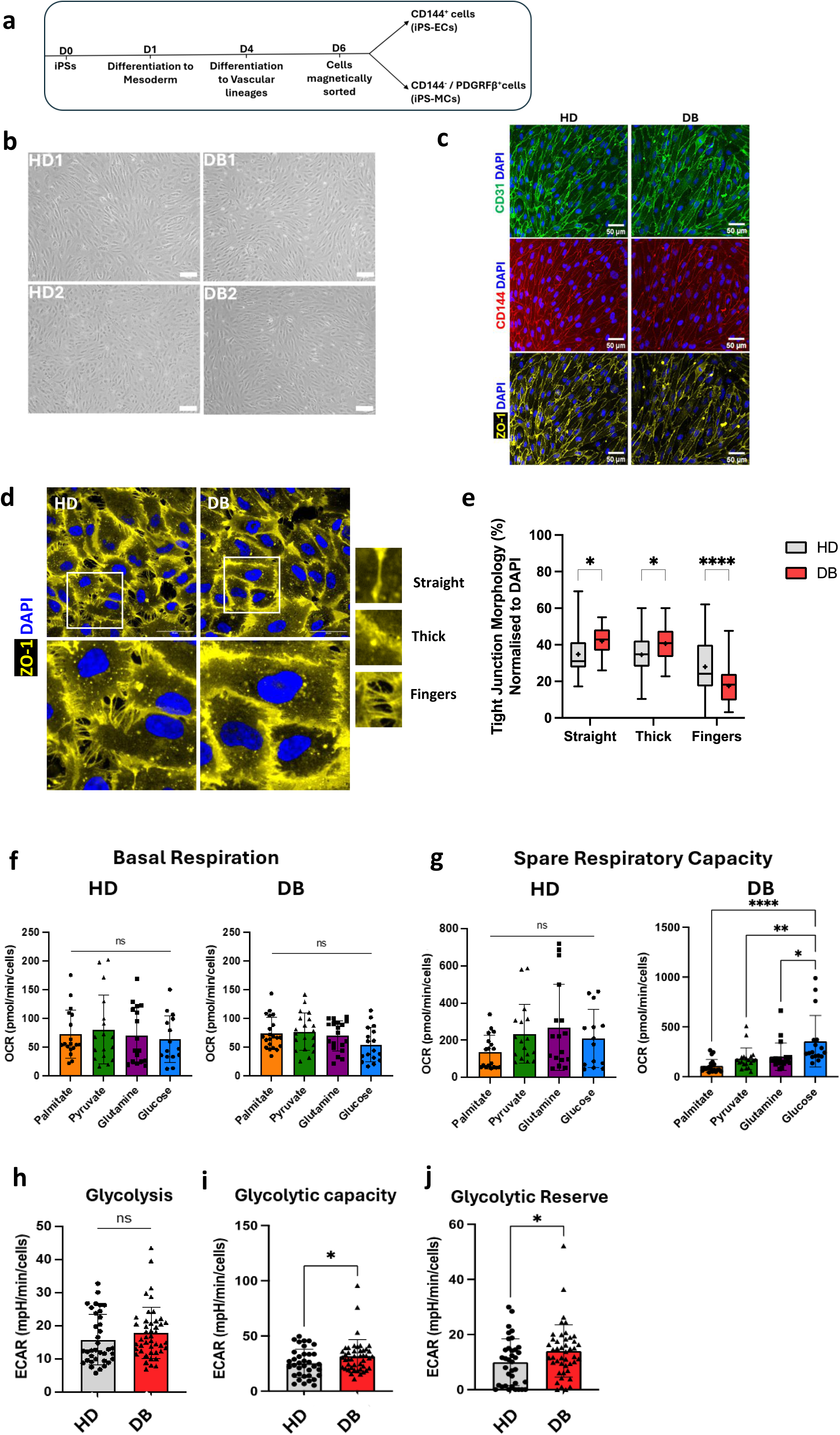
Patient iPS derived Vascular Cells: Characterization of iPS-Endothelial Cells (iPS-ECs). **(a)** Timeline of differentiation of iPS to vascular cells and magnetic isolation of CD144^+^ and CD144^-^/PDGFRβ^+^ cells. **(b)** Brightfield images of patient iPS-derived ECs (iPS-ECs) generated from healthy-donor (non-diabetic, HD) (n=2) and diabetic-donor (DB) (n=2) iPS lines displaying typical endothelial morphology. **(c)** Immunofluorescence imaging of iPS-ECs demonstrating EC-specific markers CD31 (green), CD144 (red), ZO-1 (yellow) co-stained with DAPI (blue). **(d)** Phenotypic characterization of tight junction morphology using ZO-1 (yellow) co-stained with DAPI (blue). Right insets show each type of tight junction: straight, tight, and fingers. **(e)** Quantification of tight junction morphology. Values presented as Min-Max, + indicates mean value; P-values calculated using two-way ANOVA with Bonferroni multiple comparisons test. Values presented as mean±SD; P-values calculated using two-tailed Student’s t-test. (*p<0.05, ****p<0.0001). Scale bars 50µm. **(f)** Quantification of Oxygen Consumption Rate (OCR) in iPS-ECs provided with palmitate, pyruvate, glutamine, glucose to measure basal respiration and **(g)** spare respiratory capacity in HD and DB iPS-ECs as measured using mito stress test on a Seahorse XFe24 analyser. n=3 independent preparations per line and n=3 wells per substrate. Values presented as mean±SD; P values calculated using one-way ANOVA with Bonferroni comparisons test. **(h)** Quantification of Extracellular Acidification Rate (ECAR) in HD and DB iPS-ECs measuring glycolysis, **(i)** glycolytic capacity, **(j)** glycolytic reserve as measured using glycolytic stress test on a Seahorse XFe24 analyser. n=3 independent preparations per line and n=5 wells per substrate. Patient iPS-derived ECs were generated from two healthy (non-diabetic) (HD) and two diabetic (DB) iPS lines. Values presented as mean±SD; P-values calculated using two-tailed Student’s t-test. (*p<0.05; **p<0.01; ***p<0.001; ****p<0.0001. ns, not significant.

### Patient-derived vascular cells and energy substrate utilization

The metabolic profile of iPS-ECs is a major determinant of their angiogenic potential. To gain insights into cell metabolism and mitochondrial function, we evaluated differences in substrate oxidation between iPS-ECs from healthy (non-diabetic) and diabetic donors, and we assessed the oxygen consumption rate (OCR) on a Seahorse Extracellular Flux Analyzer instrument^13^. While similar levels of basal respiration for all substrates were observed (Fig. 2f), the spare respiratory capacity for glucose was higher compared to palmitate and pyruvate in diabetic-donor cells versus the healthy (non-diabetic) cells (Fig. 2g). These findings indicate a higher preference in diabetic-donor iPS-ECs for glucose mitochondrial oxidation, implying the presence of a metabolic memory that primes diabetic donor cells towards preferential glucose utilization.

To compare the reliance of healthy (non-diabetic) and diabetic donor iPS-ECs on glycolysis for energy production, we quantified the extracellular acidification rate (ECAR), as an indication of lactate production. We detected similar levels of glycolysis, but significantly higher glycolytic capacity and glycolytic reserve in the diabetic-donor iPS-ECs (Fig. 2h-j). Furthermore, to assess mitochondria dynamics and analyse mitochondrial networks a mitotracker probe was used in iPS-ECs. Alterations in mitochondria networks with lower number of branches, end-point voxels, junction voxels and slab voxels in diabetic-donor cells compared to healthy (non-diabetic) donor cells were detected (Supplementary Fig. 1a-e). These findings highlight changes in mitochondria structure that can have functional implications^14^.

Endothelial and mural cells are the two integral cellular components of the microvasculature. Their interactions are pivotal in microvascular integrity. Differentiation of iPS to vascular cells also generated CD144^-^ cells that expressed PDGFRβ, NG2 and αSMA, –key markers of mural cells. Hence these cells were labelled as iPS-Mural cells (iPS-MCs) (Fig. 3a-c) and studied further. Interestingly, diabetic donor iPS-MCs demonstrated higher expression of αSMA (Fig. 3d-f) although no difference in numbers of PDGFRβ^+^/ αSMA^+^ double positive cells was observed. Substrate utilisation assessment using Seahorse assays indicated that iPS-MCs from all lines have increased spare respiratory capacity for glucose compared to other substrates (Fig. 3g, h). However, in contrast to iPS-ECs, no difference in substrate preference was observed between healthy (non-diabetic) and diabetic-donor derived iPS-MCs. Similarly, no differences in mitochondria dynamics were detected (data not shown). Nevertheless, as in iPS-ECs, diabetic-donor iPS-MCs displayed significantly higher glycolytic capacity compared to healthy (non-diabetic) donor iPS-MCs (Fig. 3i-k).

**Figure 3.**
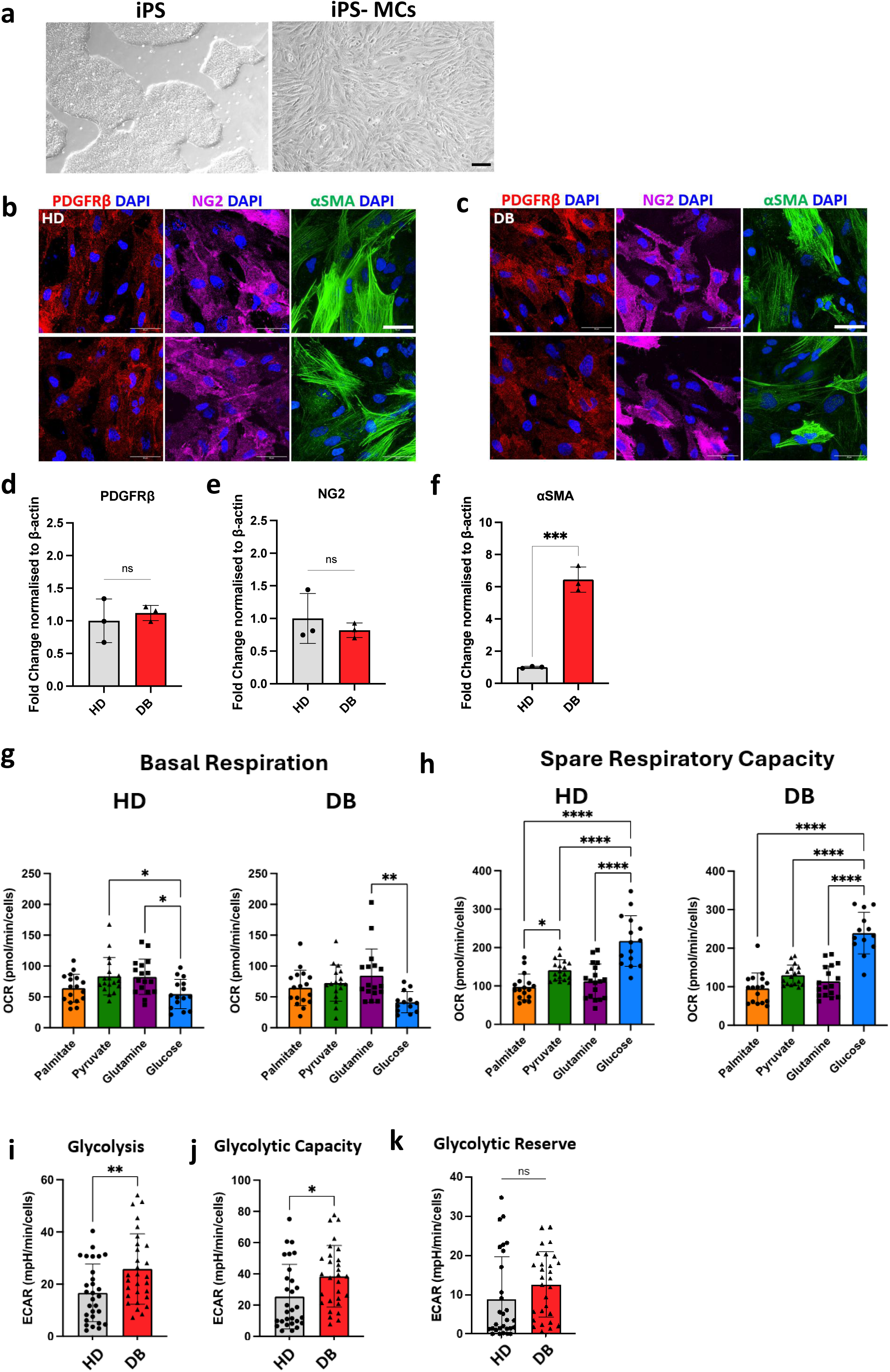
iPS-MCs Phenotypic characterization, Metabolism, Substrate Utilization, Glycolysis. **(a)** Brightfield images showing typical morphology of iPS colonies (left) and iPS-derived MCs (iPS-MCs, right). **(b, c)** Immunofluorescence imaging of iPS-MCs demonstrating MC-specific markers PDGFRβ (red), NG2 (magenta), αSMA (green) co-stained with DAPI (blue). **(d)** qRT-PCR quantification of PDGFRβ, **(e)** NG2, **(f)** αSMA in HD vs DB iPS-MCs. Values presented as mean±SD; P-values calculated using two-tailed Student’s t-test. **(g)** Quantification of OCR-linked basal respiration, **(h)** spare respiratory capacity in iPS-MCs provided with palmitate, pyruvate, glutamine, glucose. n=3 independent preparations per line and n=3 wells per substrate. Values presented as mean±SD; P values calculated using one-way ANOVA with Bonferroni comparisons test. **(i)** Quantification of ECAR in HD and DB iPS-ECs measuring glycolysis, **(j)** glycolytic capacity, **(k)** glycolytic reserve. n=3 independent preparations per line and n=5 wells per substrate. Patient iPS-derived MCs were generated from two healthy (non-diabetic) (HD) and two diabetic (DB) iPS lines. Values presented as mean±SD; P-values calculated using two-tailed Student’s t-test. (*p<0.05; **p<0.01; ***p<0.001; ****p<0.0001. ns, not significant. Scale bars 50µm.

### Diabetic media treatment of patient BVOs

Using the reference iPS cell line KOLF2, we demonstrated that maintaining BVOs in diabetic media for 7 days triggered disruption of the EC-PC crosstalk, loss of PC coverage and thickening of the capillary basement membrane (Fig. 1). These are all functional features also readily observed in diabetic clinical microvascular specimens. To determine the translational potential of the system, we used patient-derived iPS lines to generate BVOs. Following 7-day diabetic media treatment patient-derived BVOs displayed similar remodelling to the reference iPS line, KOLF2 (Fig. 4). PC dropout from the microvessels (Fig. 4f), accompanied by increased deposition of ColIV was observed in diabetic media treated BVOs derived from both diabetic-donor and healthy (non-diabetic) donor iPS lines (Fig. 4g-i).

**Figure 4.**
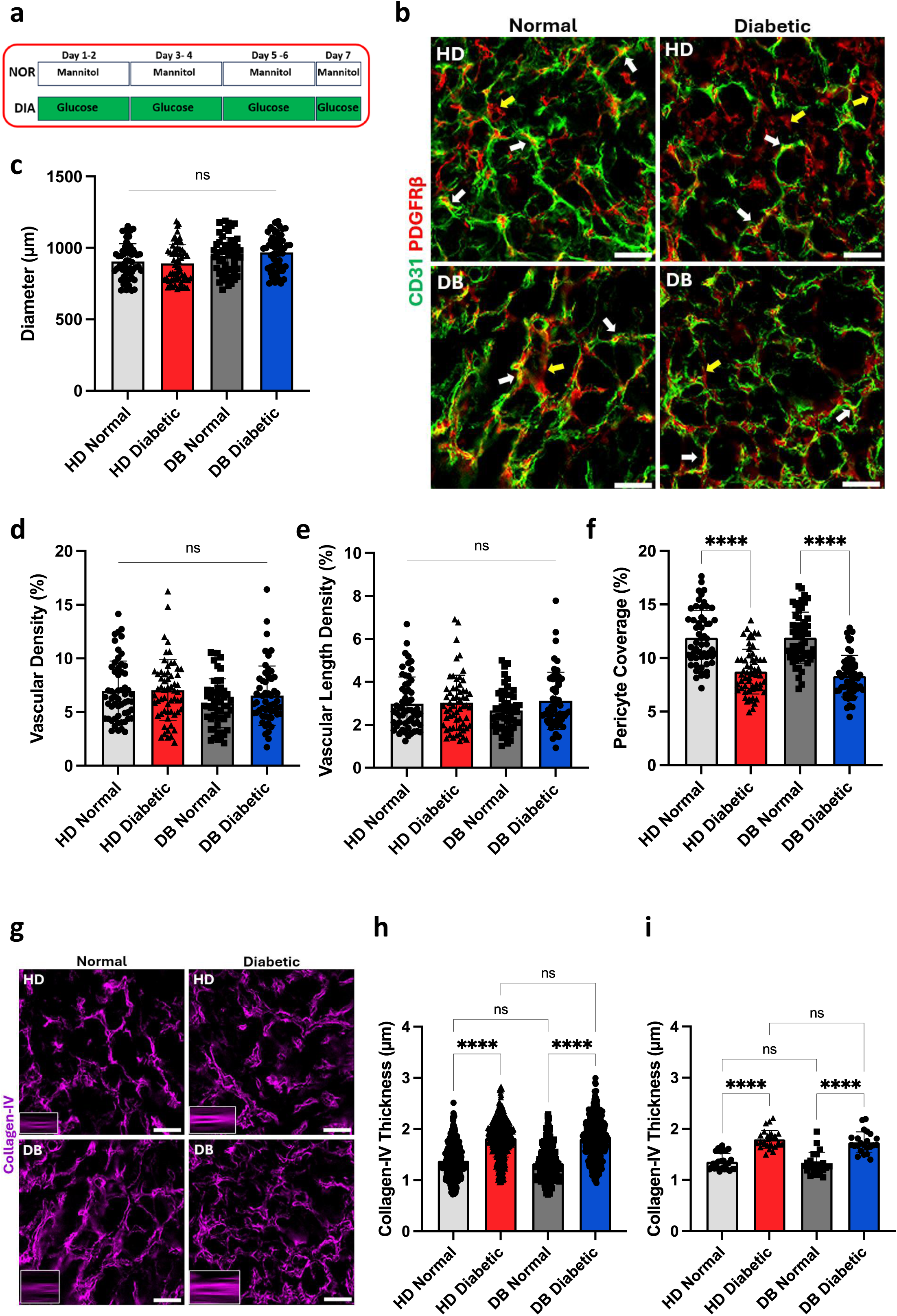
Patient-derived BVOs and treatment with diabetic media. **(a)** Schematic summarizing the media treatment with either mannitol (‘normal media’) or glucose+TNFα+IL6 (‘diabetic media’) for 7 days. Fresh media was provided every other day. **(b)** Immunofluorescence imaging of BVO sections from healthy (HD) and diabetic (DB) donor derived BVOs showing EC vessels (CD31, green) and PCs (PDGFRβ, red) in both normal and diabetic media treatment groups. White arrows indicate PCs attached to microvessels and yellow arrows indicate extravascular mural cells. **(c)** Quantification of BVO diameter from n=10-16 BVOs across 3 independent experiments per line. One-two sections per BVO were assessed; **(d)** vascular density, and **(e)** vascular length density from N=10-16 BVOs across 3 independent experiments per line in three areas per 10x image. **(f)** Percentage of pericyte coverage from n=10-16 BVOs across 3 independent experiments per line. One-two sections per BVO were assessed. **(g)** Immunofluorescence imaging of BVO sections showing collagen-IV deposition (magenta). Insets show vessel cross sections. **(h)** Quantification of collagen-IV thickness per cross-section and **(i)** per BVO section measured by re-slicing z-stacks on the vertical axis where n=150-250 cross-sections quantified from 10-16 hBVOs across 3 independent experiments per line. Patient-derived BVOs were generated from two healthy (non-diabetic) (HD) and two diabetic (DB) iPS lines. Values presented as mean±SD; P-values calculated using one-way ANOVA with Bonferroni post hoc test. ****p<0.0001. ns, not significant. Scale bars 50µm.

### Diabetic BVO microenvironment displays a distinct composition of ECM and soluble factors

An in-depth understanding of the molecular signature of diabetic microangiopathy is essential for designing tailored therapies to alter the disease trajectory. The observed microvascular remodelling integrates alterations mediated by both direct cell-cell interactions^15^ and paracrine signalling^16^. Therefore, to identify the emerging critical effectors of the microangiopathy, we generated BVOs from KOLF2 iPS cells and performed proteomic analysis of BVOs and the BVO secretome following normal and diabetic media treatment. Regulators of angiogenesis, cell-cell adhesion and cell migration displayed pronounced changes in the diabetic media treated BVOs (Fig.5a, b). This was also accompanied by alterations in actin cytoskeleton and actin binding, suggesting important changes in cell migration. Additionally, extensive ECM reorganization with remodelling of the basement membrane was detected in diabetic BVOs, highlighting the structural alterations in the diabetic microenvironment. Proteomic analysis of the BVO secretome revealed a new profile of ECM components and secreted angiogenic factors in the conditioned media that differ in normoglycemic and diabetic media treated BVOs (Fig. 5c, d).

**Figure 5.**
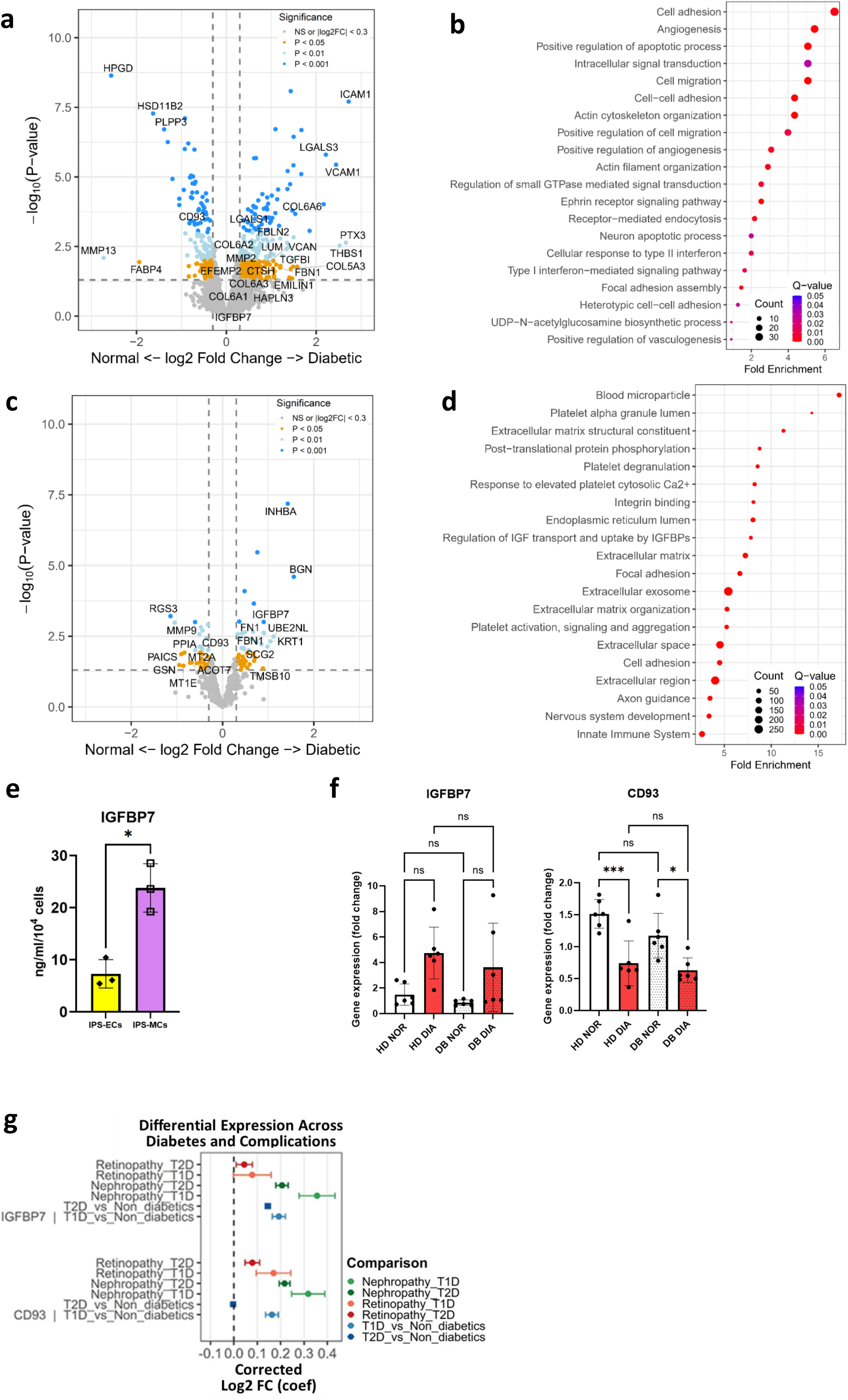
Diabetic BVO microenvironment displays a distinct composition of ECM and soluble factors. **(a)** Volcano plot comparing the proteome in the Diabetic media treated and Normoglycemic BVOs groups. **(b)** Pathway enrichment of proteins detected in the diabetic media treated BVOs. n=5 BVOs per pooled sample, from 3 separate preparations. **(c)** Volcano plot comparing the secreted proteins in conditioned media in the two groups. **(d)** Pathway enrichment of proteins detected in the diabetic BVOs. n=8 BVOs per pooled secretome sample, from 3 separate preparations. **(e)** Quantification of IGFBP7 in the conditioned media of iPS-ECs and iPS-MCs within 24h at baseline conditions. n=3 independent differentiation experiments. The KOLF2 iPS line was used to generate the BVOs in **(a-d)** and iPS-ECs, iPS-MCs **(e)**. **(f)** IGFBP7 and CD93 gene expression as assessed by qRT-PCR, β-tubulin was used as a normalization control. Patient-derived BVOs were generated from two healthy (non-diabetic) (HD) and two diabetic (DB) iPS lines. n=6 BVOs per group, (*p<0.05; ***p<0.001). ns, not significant. **(g)** Forest plot presenting the results of differential expression analysis of IGFBP7 and CD93 between Retinopathy vs Control and Nephropathy vs Control in T1D, T2D and control participants of UK Biobank and between T1D patients vs non-diabetics and T2D and non-diabetics. Analysis was done using Ebayes method of the limma package correcting for Age and Sex status.

### Increased levels of IGFBP7 protein in the BVO secretome in microvascular dysfunction

High levels of Insulin-like growth factor binding protein-7 (IGFBP7) protein were detected in the diabetic BVO conditioned media (Fig. 5c). IGFBP7 was also increased in the secretome of PFKFB3 knockout BVOs (a model of microvascular dysfunction that we previously reported^5^) compared to their isogenic control (Supplementary Fig.S2). These findings in two models of microvascular dysfunction (diabetic media treatment and metabolic rewiring through glycolysis inhibition^5^) indicate that IGFBP7 is robustly associated with microangiopathy.

IGFBP7 was previously linked to adverse clinical outcomes in HF patients and is considered a marker of cellular ageing^17^. In the heart both ECs and cardiomyocytes have been proposed as the main sources of IGFBP7^7,18^ while in human scRNAseq datasets from various organs a robust expression of IGFBP7 in mural cells and ECs is detected^19–21^ (Supplementary Fig. S3). In our system, in iPS-derived vascular cells, IGFBP7 is secreted by both cell types, with mural cells demonstrating the highest levels of IGFBP7 (Fig. 5e).

The glycoprotein CD93 plays an important role in the regulation of the angiogenic process. Expressed predominantly in ECs, CD93 acts as a soluble growth factor and an adhesion molecule^8^. In the secretome of diabetic BVOs, a decrease of soluble CD93 was observed (Fig. 5c). Recently, IGFBP7 has been identified as a ligand for CD93^8^. In our system, qPCR quantification in healthy (non-diabetic) and diabetic-donor BVOs revealed reduced expression of CD93 in diabetic media treated BVOs for all patient-derived lines (Fig. 5f). These results were highly reproducible across six patient-derived iPS lines and the reference iPS line KOLF2 (data not shown). These findings indicate that in the diabetic microvasculature the CD93-IGFBP7 signalling axis is dysregulated.

In a clinical setting, we interrogated the link between IGFBP7 and CD93 in microangiopathy patients in the UK Biobank, a large prospective study and biomedical database with information on biological samples from half a million UK participants. Data from people living with type 1 (T1D) and type 2 (T2D)^22,23^ diabetes with nephropathy and/or retinopathy, two key microvascular complications and non-diabetics were used for this analysis. Since only patients with OLINK Explore Proteomics measurements (Supplementary Fig.4) could be used, this analysis was conducted on 611 T1D patients (231 with retinopathy and 286 with nephropathy), 4944 T2D patients (729 with retinopathy and 1621 with nephropathy) and 47550 people without diabetes (1965 with retinopathy and 5393 with nephropathy). IGFBP7 was increased in the circulation in both T1D and T2D diabetics compared to non-diabetics and was further elevated in T1D and T2D nephropathy but not in retinopathy. On the other hand, increased CD93 levels were detected for both T1D and T2D nephropathy and retinopathy (Fig. 5g).

### Genetic deletion of IGFBP7 triggers ECM remodelling in BVOs

To obtain mechanistic insights into the role of IGFBP7 in the microvasculature, we generated IGFBP7 knockout iPS cells (KO-IG7) using CRISPR-Cas9 gene editing (Supplementary Fig.S5). Proteomic analysis of KO-IG7 and control BVOs demonstrated that IGFBP7 plays a pivotal role in the vasculature (Fig. 6a). Differential expression of proteins involved in adherens junctions (CDH5, DSP, EFNB2, TBCD, TJP), focal adhesions (ACTN1, VCL, ZYX), ECM structure (ACAN, COL1A2, COL3A1, COL5A1, COL6A1, COL6A2, COL6A3, DCN, FBN1, LUM) and cell-ECM binding (ADAM9, INGA2, ITGA3, ITGAV, ITGB3, SPARC) was observed. Silencing IGFBP7 in BVOs also led to profound reorganization of the actin cytoskeleton with alteration in actin filament bundle assembly (CALD1, EZR, LCP1, LIMA1, MICAL1, PDLIM1) and actomyosin structure organization (ACTC1, ACTG1, CFL2, LIMCH1, MYH9, TPM1). These findings indicate that IGFBP7 is critical in cell-cell adhesion and cell migration in the microvasculature (Fig. 6b). Pathway enrichment GSEA analysis also revealed, a significant increase in proteins involved in oxidative phosphorylation (Supplementary Fig. S6a, b), indicating that IGFBP7 silencing can trigger metabolic reprogramming in BVOs. This inverse correlation of IGFBP7 and oxidative phosphorylation was also previously reported in transgenic mice^24^. Hierarchical clustering of transcription factors based on their activity profiles in KO-IG7 and control BVOs indicated clearly distinct regulatory networks (Fig. 6c, Supplementary Fig. S6c).

**Figure 6.**
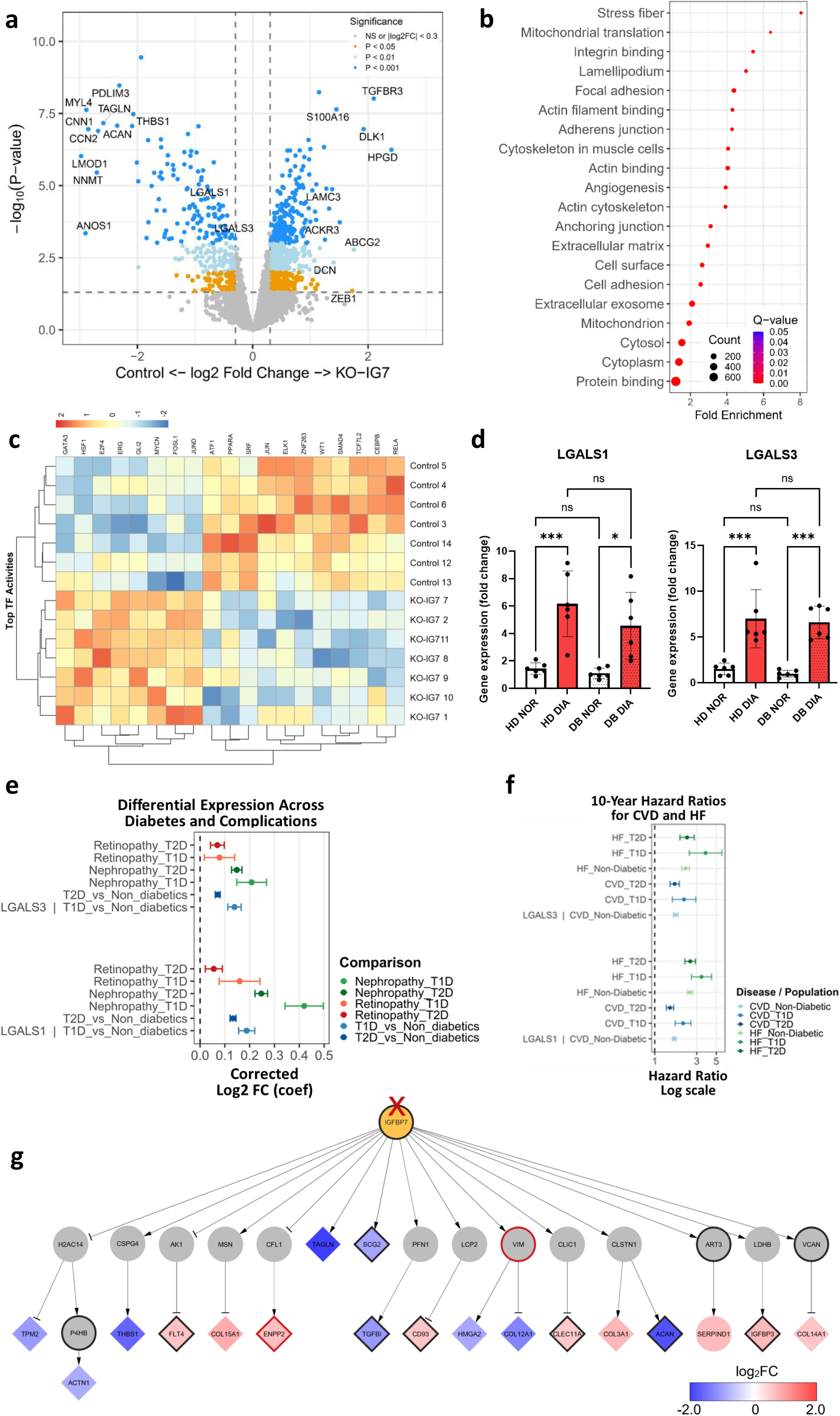
IGFBP7 regulates microvascular integrity. **(a)** Volcano plot comparing the proteome in the IGFBP7 Knockout (KO-IG7) and control BVOs. **(b)** Pathway enrichment of proteins detected in the KO-IG7 BVOs. N=7 samples per group, (9 BVOs per pooled sample), from 3 separate preparations. **(c)** Heatmap showing inferred transcription factor (TF) activity scores across KO-IG7 and control BVOs. TF activities were estimated using the VIPER algorithm with DoRothEA regulons (confidence levels A–B) implemented in the decoupleR R package. Each row represents a transcription factor, and each column represents an individual sample. Activity scores are row-scaled (z-score normalized) to emphasize relative changes in TF activity across conditions. TFs were hierarchically clustered based on similarity of activity profiles, while samples were ordered by experimental condition (control followed by KO-IG7). The KOLF2 iPS line was used to generate the BVOs in **(a-c)**. **(d)** LGALS1 and LGALS3 gene expression as assessed by qRT-PCR, β-tubulin was used as a normalization control. N=6 BVOs per group. Patient-derived BVOs were generated from two healthy (non-diabetic) (HD) and two diabetic (DB) iPS lines. N=6 BVOs per group. Data are shown as mean ± SD using one-way ANOVA followed by Tukey’s multiple comparisons test. (*p<0.05; ***p<0.001). ns, not significant. **(e)** Forest plot presenting the results of differential expression analysis of LGALS1 and LGALS3 between Retinopathy vs Control and Nephropathy vs Control in T1D, T2D and control participants of UK Biobank and between T1D patients vs non-diabetics and T2D and non-diabetics. Analysis was done using Ebayes method of the limma package correcting for Age and Sex status. **(f)** Forest plot presenting Hazard Ratio’s and 95% confidence intervals of LGALS1 and LGALS3 in prognosing 10-year CVD and HF outcomes in T1D, T2D and non-diabetic participants of UKB. Analysis was corrected for Age and Sex, while patients with pre-existing CVD diagnosis were excluded. **(g)** Inferred regulatory network integrating the diabetic BVO and PFKFB3 knockout BVO secretome datasets, showing paths from IGFBP7 to the top 10 upregulated and 10 downregulated proteins in KO-IG7 versus control BVOs. Node colour gradient indicates log2 fold change, and diamond-shaped nodes represent significantly differentially expressed proteins. Node border colour represents UK Biobank microvascular associations (black: T1D and T2D; red: T2D only). Edge arrows indicate inferred regulatory effects, with edge width proportional to interaction strength.

Of particular interest, in the KO-IG7 BVO proteome there was a downregulation of LGALS1 and LGALS3 (Fig. 6a). These glycan-binding proteins that can interact with cell surface glycans and glycosylated matricellular components, mediate protein-protein and protein-glycan binding and modulate cell-cell communication and cell-matrix interactions^25^. In the cardiovascular system, LGALS1 and LGALS3 function as mediators of neovascularization by facilitating plasma membrane retention and phosphorylation of the VEGFR2^26^. In our model, LGALS1 and LGALS3 expression levels increased in hyperglycemic-media treated BVOs, in both healthy (non-diabetic) and diabetic-donor derived BVOs (Fig. 6d). These results were highly reproducible across six patient-derived iPS lines and the reference iPS line KOLF2 (data not shown).

In clinical specimens, data from the UKB indicated that the increase in IGFBP7 in T2D and T1D individuals coincides with a higher expression of LGALS1 and LGALS3 in patients with retinopathy and people with nephropathy (Fig. 6e). The increase was more pronounced in individuals with T1D nephropathy particularly for LGALS1 levels. In terms of the increased 10-year hazard ratio for CVD and HF, both LGALS1 and LGALS3 demonstrated increased association with adverse events, in all groups (T1D, T2D and non-diabetics) with the biggest variability detected in T1D individuals (Fig. 6f).

Finally, a list of eQTls and clinically relevant variants has been identified for the IGFBP7-CD93-LGALS1-LGALS3 regulatory axis (Supplementary Table 1). Theses variants were further analysed in the All of Us (AoU) cohort to associate them with prevalent and ever diagnosed diabetes and diabetic microangiopathy complications and with future cardiometabolic outcomes (Supplementary Fig. S7, S8). A panel of 8 variants were significantly (p-value<0.05) associated with prevalent diabetes and diabetic complication diagnosis, 11 variants were significantly associated with ever-diagnosed diabetes and diabetic complication diagnosis, and 10 variants were significantly associated with at least one cardiometabolic future outcome. As shown in Supplementary Fig. S7, the variant with the highest effect size was rs570122247 a likely benign (according to ClinVar annotation), missense variant of IGFBP7 which was significantly more frequent in T2D patients and significantly associated with elevated risk of developing T2D in the next 5 years. In opposite, rs181337247, another likely benign but synonymous variant in LGALS3, was negatively associated with the frequency of T1D and T2D diagnosis.

Inferred regulatory network analysis integrating the diabetic media treated BVO and PFKFB3 knockout BVO secretome datasets with differential expression data from IGFBP7 knockout BVOs (Fig. 6g) suggested that IGFBP7 may regulate microvascular integrity through downstream interactions involving extracellular matrix proteins (THBS1 and TGFBI), collagen proteins (COL15A1, COL3A1, and COL14A1), angiogenic regulators (FLT4, CD93, and ENPP2), and cytoskeletal proteins including TPM2 and ACTN1. Several downstream proteins, including IGFBP3, CLEC11A, and ACAN, were additionally associated with diabetic microvascular complications in UKB analyses.

## Discussion

Human organoids that share functional and histological characteristics with their organ counterparts are increasingly used as models of biology and disease^27^. These human tissue systems could overcome the mismatch between animal and human biology that led to translational failures in the last decades^28^. Derived from patient iPS cells, organoids can also serve as a personalized platform for targeted therapies.

In this study, to establish a personalised system of diabetic microangiopathy, we used patient-derived iPS lines and a reference iPS line to generate BVOs, a model of human microvasculature^5,6^. Using treatment with media supplemented with high glucose and low cytokines for 7 days, we observed a disruption of the PC-EC crosstalk and ECM thickening in diabetic media treated BVOs. These are hallmarks of diabetic microangiopathy also observed in diabetic patient tissue specimens^2,3,6^, indicating that our model faithfully recapitulates the early features of diabetic microangiopathy. We could reproducibly generate BVOs from all patient iPS lines, suggesting that this is a robust model of disease. Metabolomic analysis of diabetic-donor and healthy (non-diabetic) donor iPS-derived iPS-ECs indicated a predisposition for the use of glucose and altered mitochondria dynamics in diabetic-donor cells. Our findings provide support to previous reports of diabetic mitochondrial impairment^29^ and pro-inflammatory priming in diabetes following infection,^30^ and may imply that patient derived BVOs could have an added value in establishing novel therapeutic approaches.

Using two human tissue models of microvascular dysfunction, namely knocking out PFKFB3 to trigger metabolic rewiring through glycolysis inhibition and diabetic media treatment of BVOs, we demonstrated a robust upregulation of IGFBP7 protein in the BVO secretome, with iPS-MCs secreting substantially higher levels compared to iPS-ECs. Based on the qRT-PCR quantification, the elevation of IGFBP7 protein in the diabetic secretome is likely the result of protein accumulation. This was similar in healthy (non-diabetic) and diabetic-donor derived BVOs. Interrogation of diabetic microangiopathy cohorts in the UKB provided additional evidence for the translational potential of the findings in diabetic media treated BVOs. A strong association of IGFBP7 with diabetic nephropathy in T1D and T2D individuals was observed.

A predictive value for IGFBP7 in the progression of the diabetic kidney disease has recently been reported. The IGFBP7 levels were elevated in both blood and urine samples in patients during the early stages of diabetes and correlated with key indicators of diabetic kidney disease^24^. Nonetheless, IGFBP7 is best known as an indicator of senescence in the context of HF^17,31^. IGFBP7 emerged as an independent and robust prognostic biomarker in patients with HF with both reduced and preserved ejection fraction^31–33^. IGFBP7 levels strongly correlated with the presence of atrial fibrillation, diabetes mellitus, NT-proBNP levels, and worse clinical status; increased number of hospitalizations and the extent of peripheral oedema^31,32^. Although the mechanism is not well understood, an effect on cell proliferation^34^, the proinflammatory cascade and fibrosis were reported^35^.

IGFBP7 is known to bind to insulin, IGF-1 and IGF-2 and inhibit their functions, albeit with much lower affinity compared to other IGFBPs. IGFBP7 binding to unoccupied IGF1R was reported to suppress downstream signaling and inhibit protein synthesis, cell growth, and survival^36^. IGF-independent interactions of IGFBP7 include binding to CD93^37^, an interaction of particular interest as it plays a pivotal role in angiogenesis through regulation of EC angiogenic potential, migration and EC-matrix interactions^8^. In our system, diabetic media treated BVOs from healthy (non-diabetic) and diabetic donors demonstrated a downregulation of CD93 expression at the transcript level that was also reflected at the protein level both as an adhesion molecule in the BVOs and as a soluble growth factor in the BVO secretome. Intriguingly, in the UKB, diabetic microangiopathy was linked to elevated circulating levels of CD93, which might reflect on the differential contribution of other organs to circulating CD93.

Here we report for the first time a direct link between IGFBP7 and LGALS1 and LGALS3, implicating the glycocalyx in microvascular integrity. The glycan binding LGALS1 and LGALS3 have both been previously reported in angiogenesis. LGALS1, encoding galectin 1, is a prototype galectin, harbouring a single conserved carbohydrate recognition domain (CRD) that recognizes glycans. Galectin 1 can function as a monomer or form non-covalent homodimers with identical CRDs^25^. Its preferential association with N-glycans on VEGFR2 was shown to act as a glycosylation-dependent circuit that can preserve the angiogenic phenotype in the absence of VEGFA in tumour cells. On the other hand, LGALS3 encodes galectin 3, a unique chimera-type β-galactoside-binding protein that contains a CRD connected to an N-terminal domain that is responsible for oligomerization of the lectin and ligand cross-linking^25^. Galectin 3 can interact with a variety of surface molecules such as CD146, LAMP-1/-2, VEGFR2, CD44, CD45 and αvβ3 integrin to regulate cell-matrix interactions. Galectin 3 promotes EC migration and capillary formation *in vitro* and enhances vascularization *in vivo,* by regulating VEGF-R2 internalization and phosphorylation. LGALS3 knockout mice display impaired angiogenesis^38^ due to accumulation of VEGF receptors in endosomal vesicles indicating that galectin 3 is required for VEGFR2 retention on the plasma membrane. Galectin 3 levels in the circulation have strong associations with diabetes,^39^ metabolic syndrome^40^ and the severity of HF^41^. Higher galectin-3 significantly correlated with a higher risk of long-term all-cause mortality in HF patients^42^.

In our model, LGALS1 and LGALS3 expression levels increased in diabetic media treated BVOs, in both healthy (non-diabetic) and diabetic-donor derived BVOs. Proteomic analysis further highlighted the elevated galectin 1 and galectin 3 protein expression in diabetic media treated BVOs. These findings from the preclinical BVO model were also evaluated in clinical specimens. Interrogation of the UKB dataset, revealed an association of IGFBP7, LGALS1 and LGALS3 with diabetic nephropathy and retinopathy.

Previously, in biopsies from patients with chronic kidney disease, a correlation between LGALS3 and IGFBP7 mRNA expression was observed^43^. On a different system, in psoriasis, a subset of skin ECs that secrete high amounts of IGFBP7 was identified. IGFBP7 secretion was linked to the destruction of the endothelial glycocalyx and immune-cell adhesion and extravasation^44^. In psoriatic capillary ECs upregulation of IGFBP7 coincided with elevated levels of LGALS1, but the potential link between the two was not investigated.

Here, using genetic engineered IGFBP7 knockout iPS to generate BVOs we provide evidence for a mechanistic link between IGFBP7, LGALS1 and LGALS3 expression. Proteomic analysis revealed a profound reduction in galectin 1 and galectin 3 proteins in the mutant BVOs compared to control, indicating an IGFBP7-LGALS1-LGALS3 axis in the microvasculature.

The effectors we have identified using BVOs as a model of microangiopathy, could have a dual role, serving as markers of disease severity and/ or potential novel therapeutic targets. The IGFBP7-LGALS1-LGALS3 regulatory panel that emerged in our study is strongly associated with adverse cardiovascular events in diabetic patients in the large clinical cohorts of UKB, although these analyses don’t allow inferring causality. Furthermore, our proteomic analysis of engineered IGFBP7 knockout BVOs revealed a panel of downstream effectors that may serve as readouts of IGFBP7 activity. Among them, our inferred network analysis already highlights a subset of regulators quantified in the UKB, some of which also showed associations with diabetic microangiopathy. These proteins that can be readily measured in the human clinical specimens, and could be assessed in small molecules/ repurposed drugs screening to evaluate the efficiency of IGFBP7 targeting.

## Conclusions

Our study indicates that BVOs are a human preclinical model that can provide unique mechanistic insights into microangiopathy. The IGFBP7-LGALS1-LGALS3 axis that we identified in diabetic media treated BVOs is strongly linked to the disease in clinical settings, providing further support to the potential of the human tissue model in the development of novel therapeutic applications.

## Acknowledgements

This study was supported by King’s BHF Centre of Research Excellence (RE/18/2/34213), (RE/24/130035), grants from the British Heart Foundation (FS/4yPhD/F/24/34214), (PG/19/56/34550) and the Diabetes UK (24/0006757). This study was enabled by access to the UK Biobank resource (application ID 98729). The authors thank all UK Biobank-enrolled individuals for their participation.

This research used data accessed through the All of Us Researcher Workbench under the project “Multimodal cross endpoint risk prediction” (by R.O. B.W. UserID).

## Author contributions

A.Za and AMS conceived the idea, A.Za and KT designed experiments and analysis. AP, LB, SA, SR, IS, ES, SHP and A.Za performed experiments. TP performed the bioinformatic analysis of the proteomic dataset and the inferred network. KT, RS and RO performed the AoU and UKB analysis. AM provided the patient iPS lines. All the authors participated in data collection and interpretation. A.Za drafted the manuscript with critical revision feedback from all other authors. All authors read and approved the final manuscript.

## Data availability

The proteomics datasets generated and analysed during the current study have been deposited to the ProteomeXchange Consortium via the PRIDE partner repository under the data set identifiers PXD079790 and PXD079768.

## Competing interests

All authors declare no competing interests.

## Figure Legends

**Supplementary Fig. S1.**
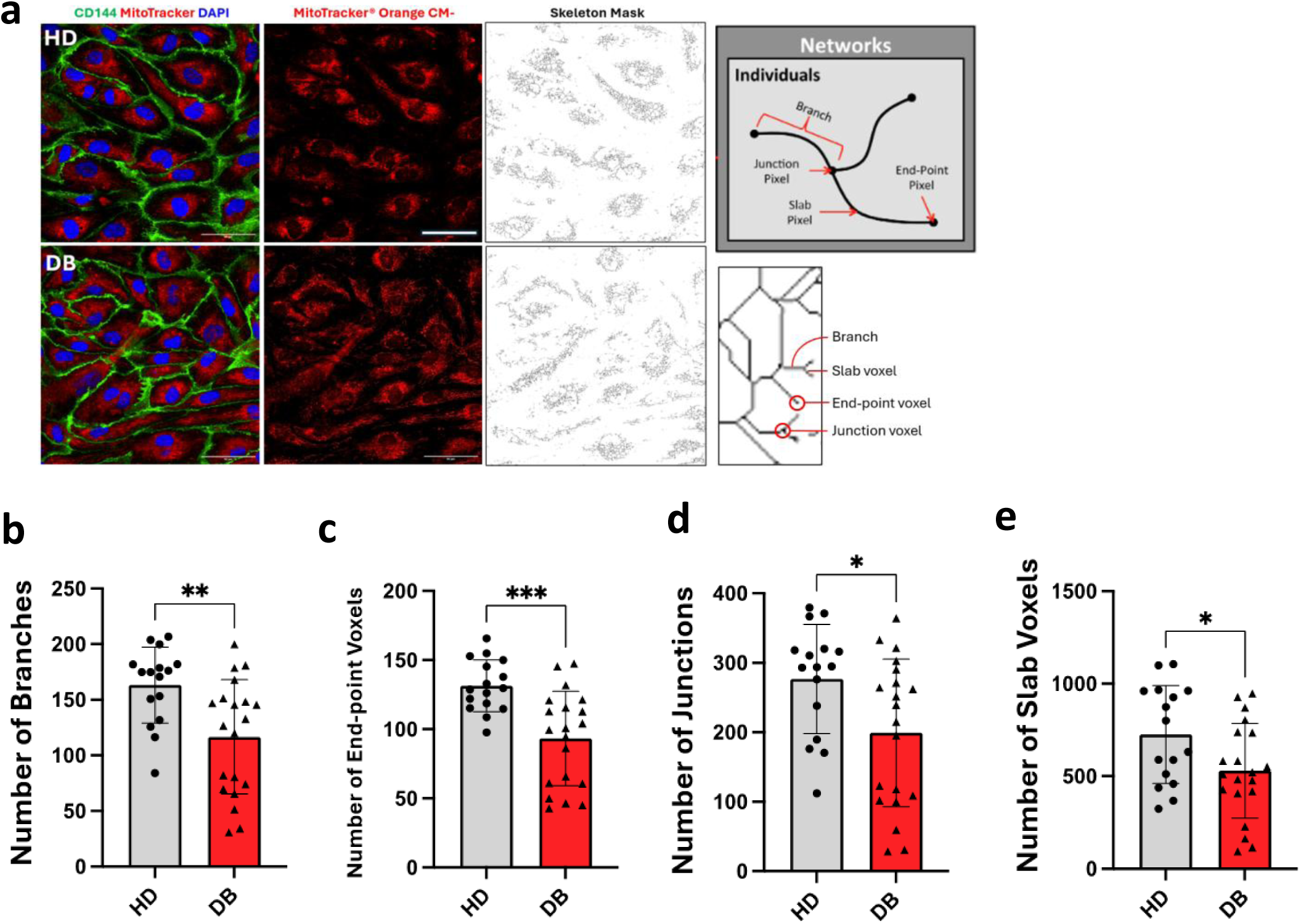
Mitochondria Dynamics in iPS-ECs. **(a)** Immunofluorescence confocal imaging of iPS-ECs with CD144 (green), MitoTracker (red), and DAPI (blue). Schematic (right) shows skeletal mask of MitoTracker with labelled mitochondrial structures. Quantification of number of **(b)** branches, **(c)** end-point voxels, **(d)** junctions and **(e)** slab voxels. Patient iPS-derived ECs were generated from two healthy (non-diabetic) (HD) and two diabetic (DB) iPS lines. Values presented as mean±SD; P-values calculated using two-tailed Student’s t-test. (*p<0.05; **p<0.01; ***p<0.001; ****p<0.0001. ns, not significant. Scale bars 50µm.

**Supplementary Fig. S2.**
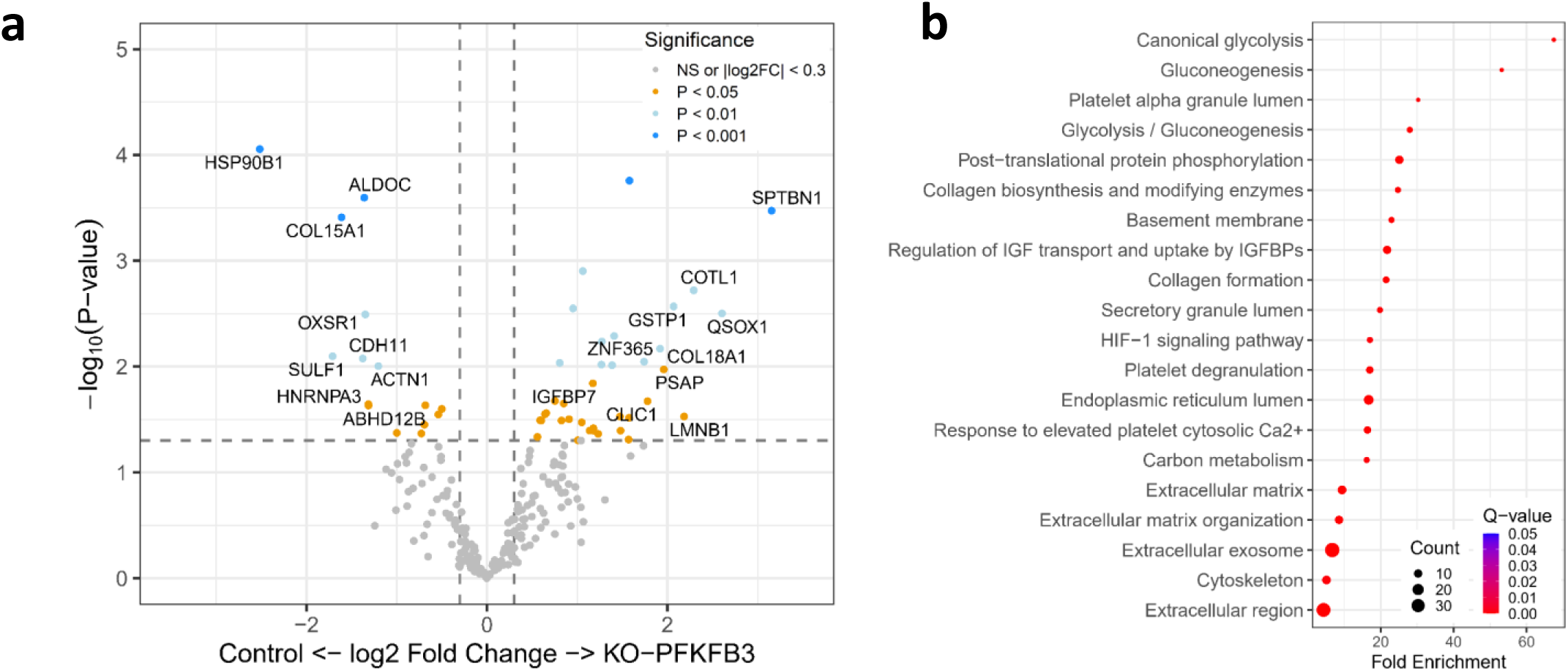
Proteomic analysis of the PFKFB3 knockout BVO secretome. **(a)** Volcano plot comparing the proteome in the PFKFB3 knockout BVOs and their isogenic control BVOs. **(b)** Pathway enrichment of proteins detected in the diabetic BVOs. n=5 BVOs per pooled sample, from 3 separate preparations. The KOLF2 iPS line was used for gene editing^5^.

**Supplementary Fig. S3.**
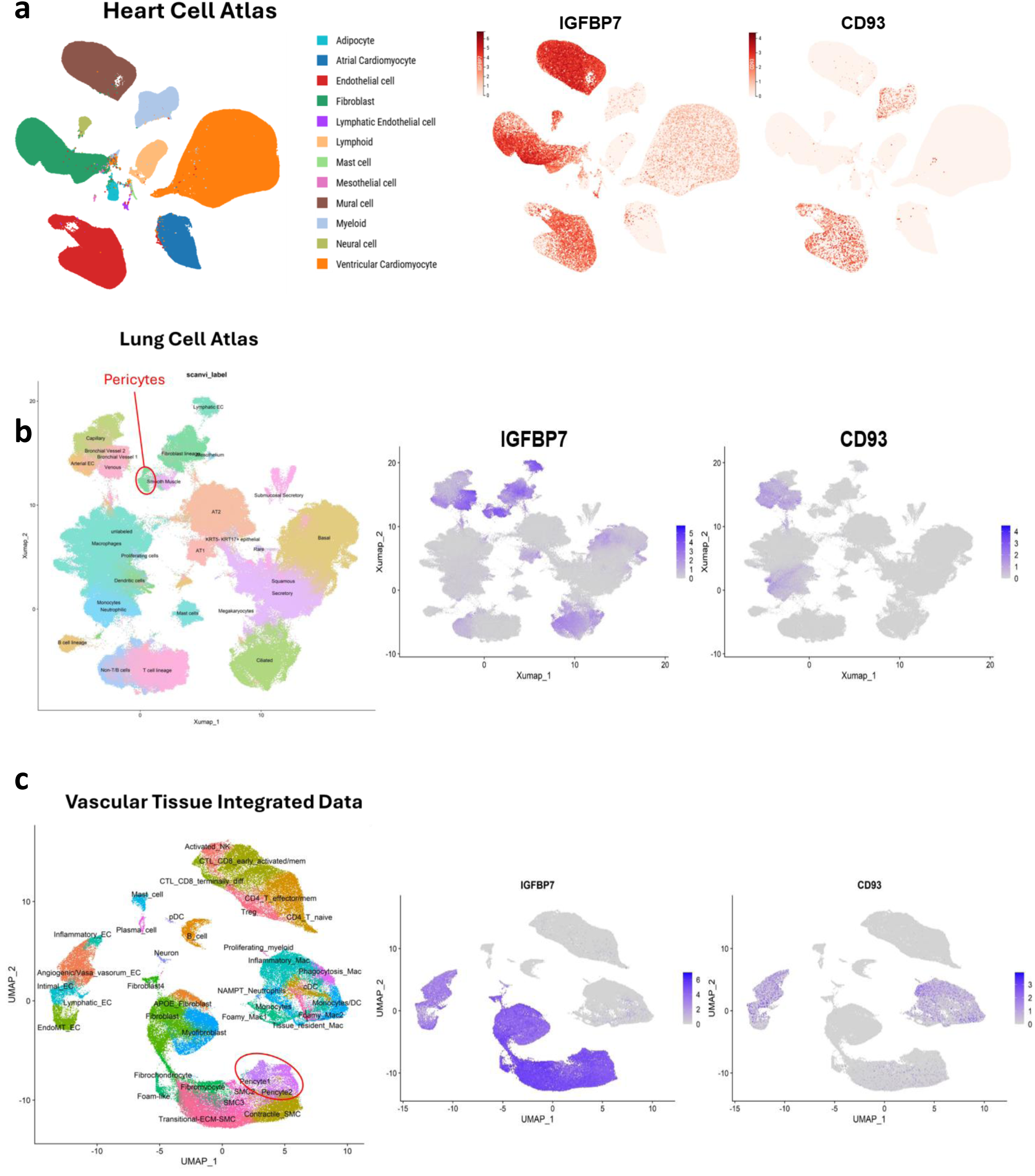
IGFBP7 and CD93 expression in human tissue scRNAseq datasets. **(a)** UMAP plot showing the expression of IGFBP7 and CD93 in Heart Cell Atlas, **(b)** UMAP plot showing the expression of IGFBP7 and CD93 in Lung Cell Atlas highlighting the pericytes cell-population, **(c)** UMAP plot showing the expression of IGFBP7 and CD93 in Mosquera et al^21^ integrated scRNAseq data from human coronary arteries and carotid plaque experiments highlighting the PCs cell-population.

**Supplementary Fig. S4.**
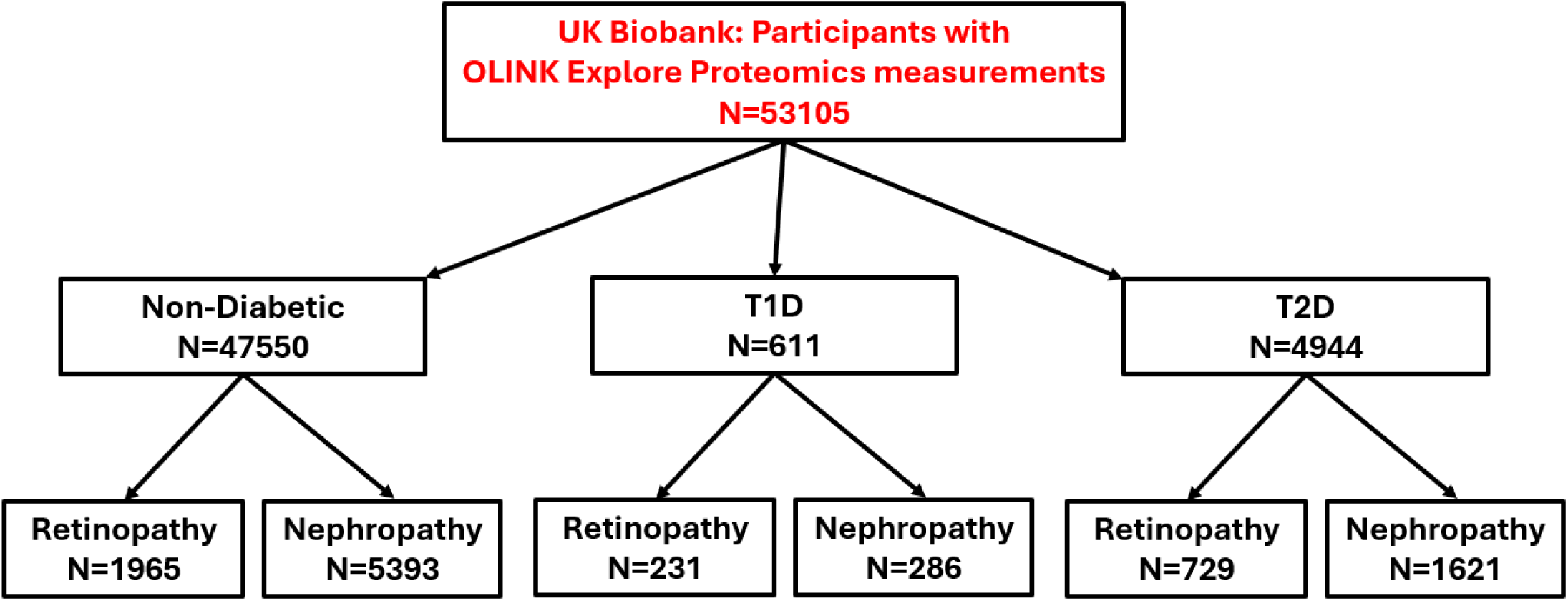
Consort Diagram of UKB participants. Patients with OLINK Explore Proteomics measurements were used this analysis.

**Supplementary Fig. S5.**
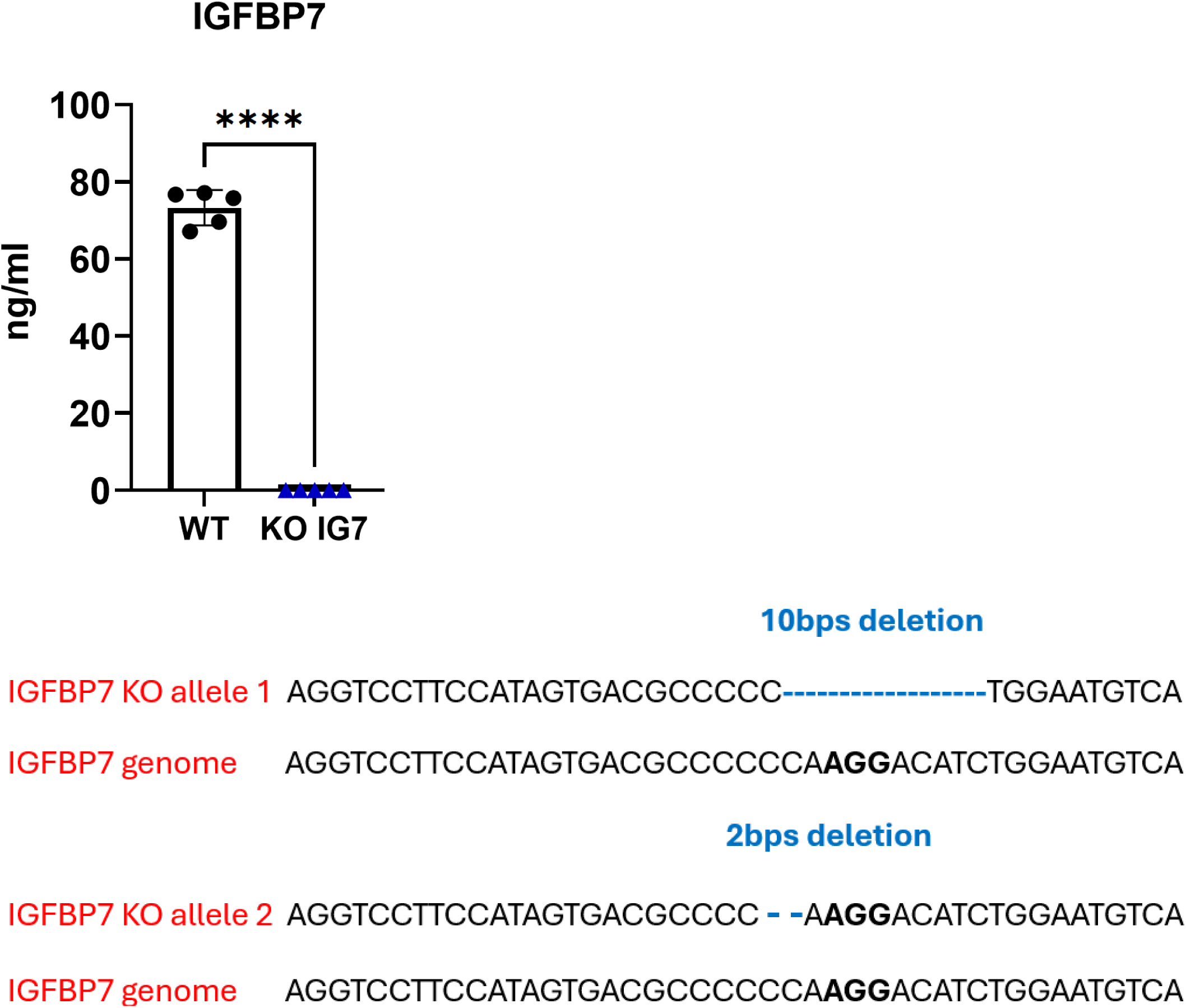
Generation of IGFBP7 knockout iPS cells. **(a)** CRISPR/Cas9 genome editing was used to generate IGFBP7 knockout iPS cells (KO-IG7) that lacked the expression of IGFBP7, as determined by ELISA on the conditioned media of BVOs. **(b)** Sanger sequencing of genomic DNA revealed the presence of deletions. The PAM sequence is highlighted in bold, deletions are shown in blue. The KOLF2 iPS line was used for gene editing.

**Supplementary Fig. S6.**
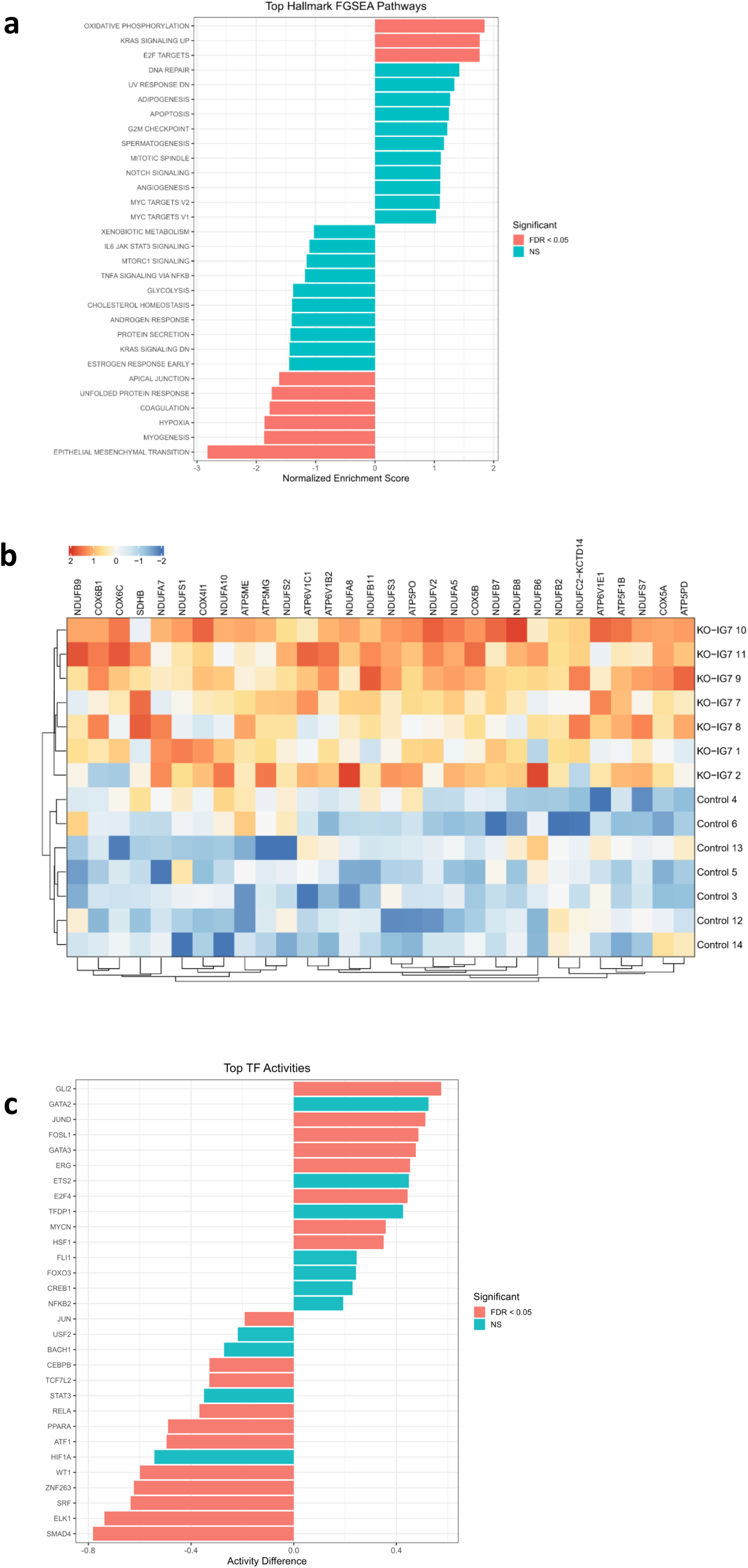
Pathway enrichment analysis for the IGFBP7 KO vs Control BVOs dataset. **(a)** Hallmark pathway enrichment analysis following IGFBP7 silencing. Fast gene set enrichment analysis (FGSEA) was performed using the Hallmark gene sets on proteins differentially expressed between IGFBP7 knockout and control BVOs identified by mass spectrometry. Bar plot shows the top 30 pathways ranked by normalized enrichment score (NES). Positive NES values indicate enrichment in IGFBP7 knockout samples, whereas negative NES values indicate enrichment in control samples. Statistically significant pathways with a Benjamini-Hochberg-corrected FDR <0.05 are coloured in red. **(b)** Heatmap of proteins from the enriched KEGG Oxidative Phosphorylation pathway in IGFBP7 knockout versus control BVOs. Heatmap showing row-wise Z-scored abundances of differentially expressed proteins belonging to the KEGG Oxidative Phosphorylation pathway, which was identified as significantly enriched in the comparison between IGFBP7 knockout (n=7) and control (n=7) pooled BVO samples. **(c)** Differential transcription factor activity between IGFBP7 knockout and control BVOs. Bar plot depicting the top 30 transcription factors exhibiting differential regulatory activity between IGFBP7 knockout (KO-IG7) and control samples. TF activity scores were inferred using VIPER-based analysis and compared between conditions using independent two-sample t-tests for each transcription factor. Bars represent the difference in mean activity (KO-IG7 − control), with positive values indicating increased TF activity in IGFBP7 knockout samples and negative values indicating decreased activity. Statistical significance was assessed using Benjamini–Hochberg corrected p-values (FDR), with transcription factors ranked by adjusted significance.

**Supplementary Fig. S7.**
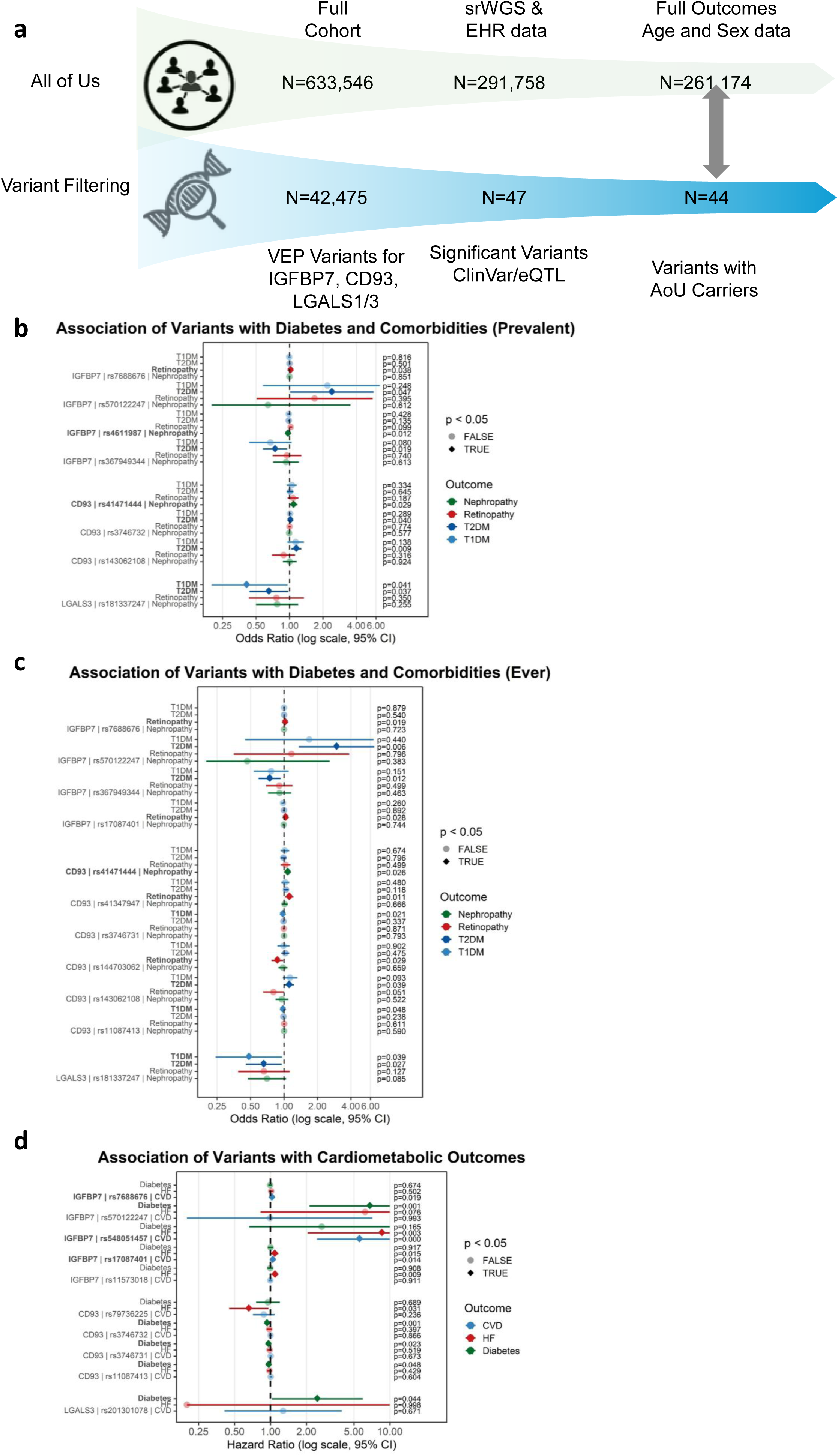
Identification of variants in diabetic microangiopathy signature in AoU cohort. **(a)** Diagram presenting the criteria for filtering All of Us patients and variants in the diabetic microangiopathy signature (IGFBP7, CD93 and LGALS1/3). (**b**) Forest plot presenting the calculated odds ratios and confidence intervals for variants significantly (p-value < 0.05) associated with at least one type of diabetes or type of diabetic complication (nephropathy and retinopathy) in the baseline of AoU or in the first 14 days. **(c)** Forest plot presenting the calculated odds ratios and confidence intervals for variants significantly (p-value < 0.05) associated with at least one type of diabetes or type of diabetic complication (nephropathy and retinopathy) at any time. **(d)** Forest plot presenting the calculated hazard ratios and confidence intervals for variants significantly (p-value < 0.05) associated with at least one type of future cardiometabolic (Diabetes, heart failure and CVD) outcome in a subpopulation of diabetes and CVD-healthy individuals at baseline.5-year follow-up was used. All analysis were corrected for Age and Sex.

**Supplementary Fig. S8.**
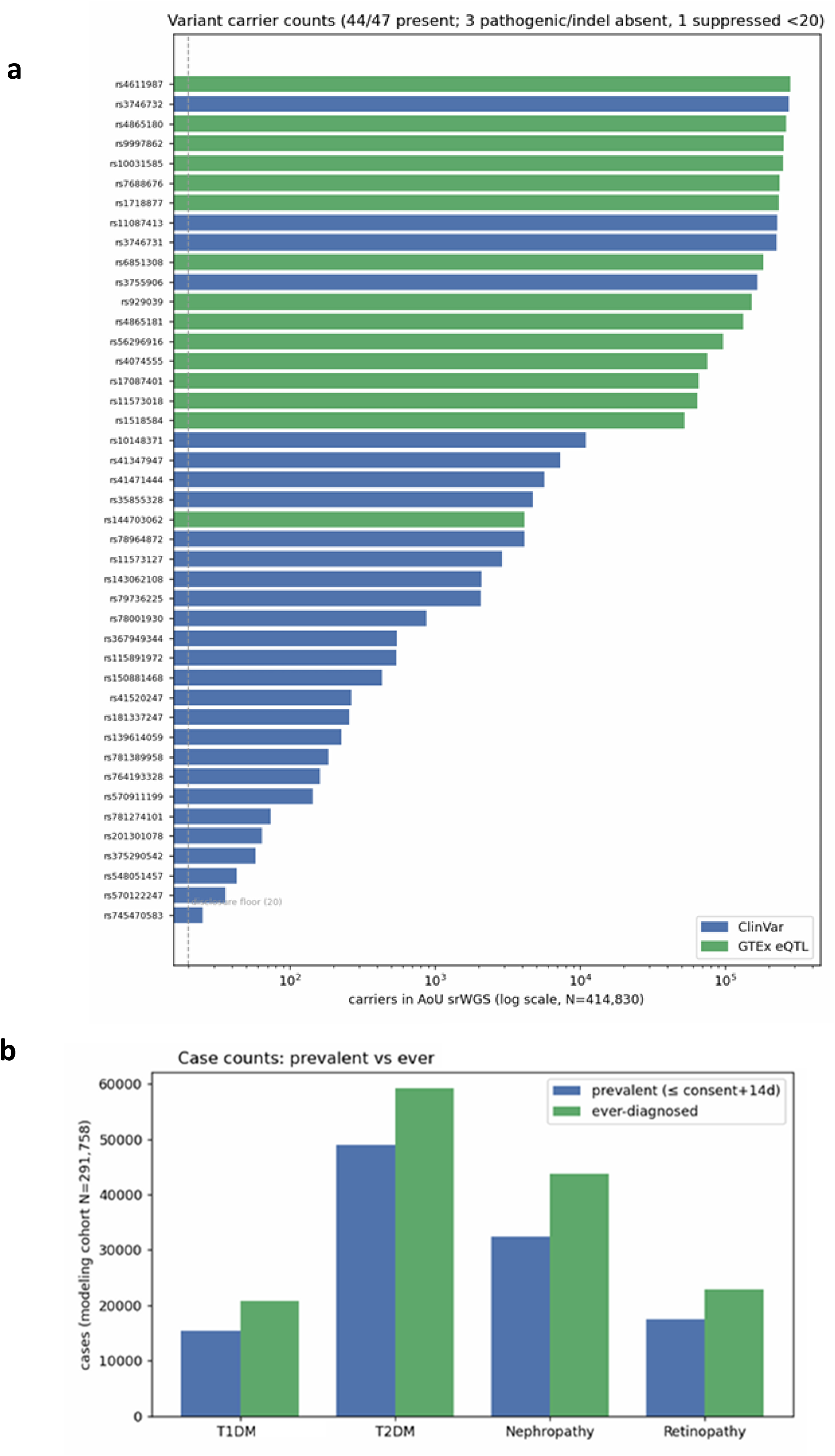
Frequencies of variants in diabetic microangiopathy signature and of diabetic endpoints in the AoU cohort. **(a)** Number of carriers of each variant in the diabetic microangiopathy signatures. **(b)** Number of case counts in each diabetes type or diabetic complication in the prevalent disease group (at presentation or within first 14 days) and the ever-diagnosed group.

**Supplementary Table 1.** The Expression Quantitative Trait Loci (eQTL) variants of IGFBP7.

**Supplementary Table 2.** Donor information for patient iPS cells used in this study.

| Study ID | Age Range (years) | Sex | BMI | Blood Pressure (mmHg) | Diabetes type | Diabetes duration |
| --- | --- | --- | --- | --- | --- | --- |
| DB1 | 71-75 | F | 29.4 | 160/70 | Type 2 | > 15 years |
| DB2 | 61-65 | M | 52.4 | 125/67 | Type 2 | > 15 years |
| DB3 | 71-75 | M | 31.3 | 148/76 | Type 2 | > 15 years |
| HD1 | 56-60 | M | 25.7 | 115/80 | Not Diabetic | N/A |
| HD2 | 56-60 | M | 28.3 | 136/83 | Not Diabetic | N/A |
| HD3 | 56-60 | M | 24.6 | 131/74 | Not Diabetic | N/A |

**Supplementary Table 3.**
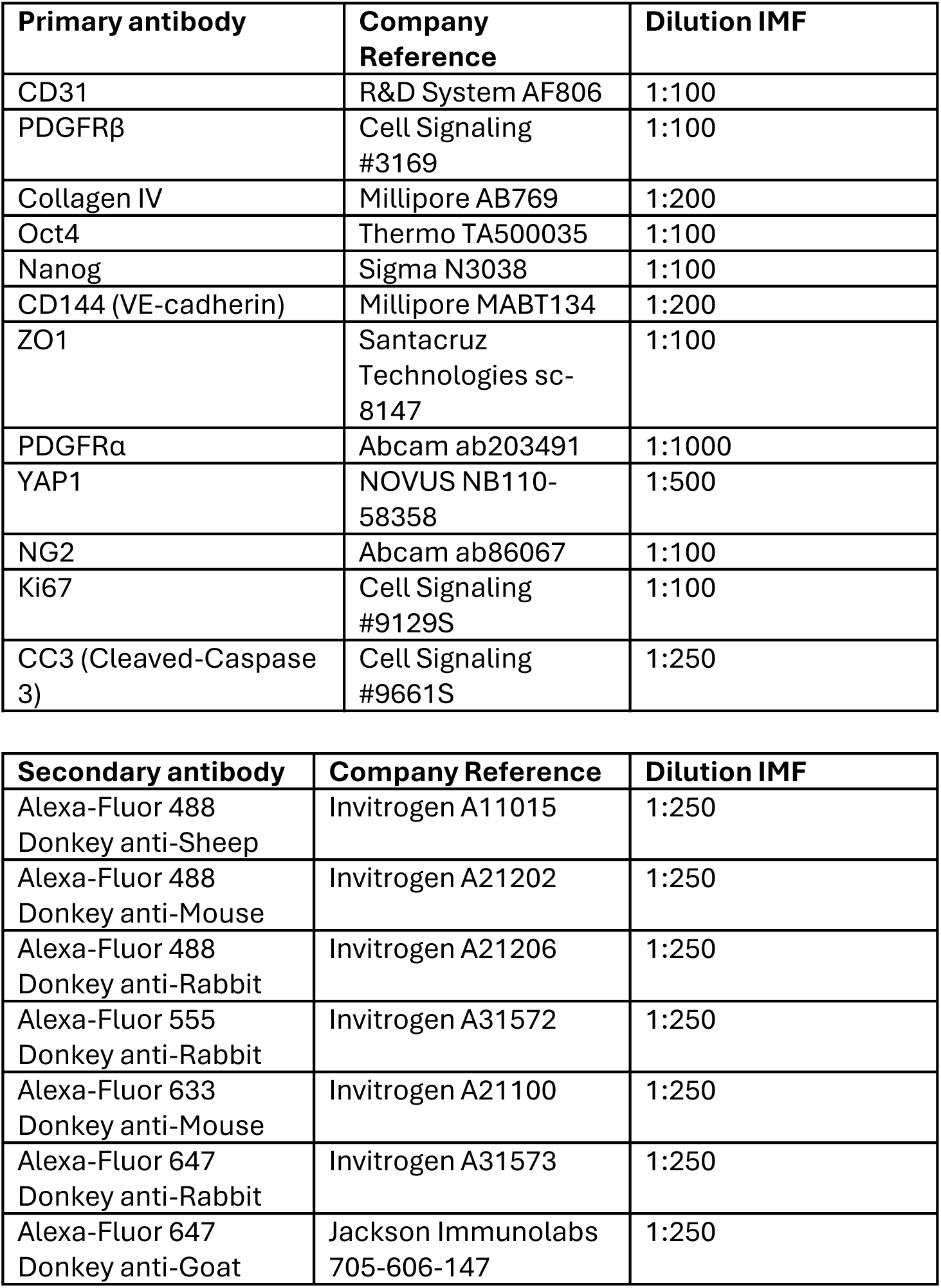
List of antibodies used for Immunofluorescence staining.

| Primary antibody | Company Reference | Dilution IMF |
| --- | --- | --- |
| CD31 | R&D System AF806 | 1:100 |
| PDGFRβ | Cell Signaling #3169 | 1:100 |
| Collagen IV | Millipore AB769 | 1:200 |
| Oct4 | Thermo TA500035 | 1:100 |
| Nanog | Sigma N3038 | 1:100 |
| CD144 (VE-cadherin) | Millipore MABT134 | 1:200 |
| ZO1 | Santacruz Technologies sc-8147 | 1:100 |
| PDGFRα | Abcam ab203491 | 1:1000 |
| YAP1 | NOVUS NB110-58358 | 1:500 |
| NG2 | Abcam ab86067 | 1:100 |
| Ki67 | Cell Signaling #9129S | 1:100 |
| CC3 (Cleaved-Caspase 3) | Cell Signaling #9661S | 1:250 |

| Secondary antibody | Company Reference | Dilution IMF |
| --- | --- | --- |
| Alexa-Fluor 488 Donkey anti-Sheep | Invitrogen A11015 | 1:250 |
| Alexa-Fluor 488 Donkey anti-Mouse | Invitrogen A21202 | 1:250 |
| Alexa-Fluor 488 Donkey anti-Rabbit | Invitrogen A21206 | 1:250 |
| Alexa-Fluor 555 Donkey anti-Rabbit | Invitrogen A31572 | 1:250 |
| Alexa-Fluor 633 Donkey anti-Mouse | Invitrogen A21100 | 1:250 |
| Alexa-Fluor 647 Donkey anti-Rabbit | Invitrogen A31573 | 1:250 |
| Alexa-Fluor 647 Donkey anti-Goat | Jackson Immunolabs 705-606-147 | 1:250 |

**Supplementary Table 4.** List of antibodies used for FACS Analysis.

| Primary antibody | Company Reference | Dilution |
| --- | --- | --- |
| CD31-AlexaFluor647 | BD Biosciences, 558094 | 5µl/test |
| CD140b-PE | BD Biosciences, 558821 | 20µl/test |
| CD144-FITC | BD Biosciences, 560874 | 20µl/test |
| CD45-FITC | Invitrogen, 11-0459-41 | 5µl(0.25µg)/test |
| CD90-PerCP/Cyanine5.5 | Biolegend, 328117 | 1µg/million cells |
| CD73-BV650 | BD Biosciences, 742633 | 10µl/test |
| CD44-PE | BD Biosciences, 550989 | 20µl/test |
| CD144-BV786 | BD Biosciences, 565672 | 5µl(0.25µg)/test |
| Live/Dead-FVS780 | BD Biosciences, 565388 | 0.1µl/test |

## Materials and Methods

### Generation of Blood Vessel Organoids

BVOs were generated from iPS cells using the protocol established by Wimmer et al, 2019^6^. The KOLF2 human iPS cell line was obtained from the Wellcome Trust Sanger Institute. The iPS lines generated from peripheral blood mononuclear cells of healthy donor (non-diabetic) (HD) and diabetic (DB) donors as described previously were also used^45^ (ethical approval from the Office for Research Ethics Committees (REC 14/NI/1109)). The HD donors exhibited normotension, body mass index (BMI)≤30 and normoglycemia, while the DB donors were people living with type 2 Diabetes mellitus (T2D) for more than 15-years^45^. Donor details are presented in the Supplementary Table 2. iPS cells were plated on Matrigel-coated (ATCC, ACS-3035) 6-well plates and cultured using the StemMACS iPS Brew-XF system (Milteney Biotech, 130-104-368). For differentiation to BVOs, 2.5×10^5^ iPS cells per well were seeded into Ultra-low adherent 6-well plates (Appleton Woods, 3471) in Aggregation Media to form cell aggregates (diameter average 50-150 µm) for approximately one day. Aggregates were collected by gravitation (Day 0) and resuspended into the mesodermal induction media (N2B27 Media supplemented with 12 μM CHIR (Tocris Bioscience, cat. no. 4423) and 30 ng/ml BMP4 (ThermoFisher, PHC9534)^6^. On day 3, the aggregates were collected by gravitation and plated in N2B27 media supplemented with 100ng/ml VEGFA (Peprotech, 100-20) and 2 μM Forskolin (R&D system, 1099) for vascular lineage promotion. On day 5, the aggregates were embedded in a substrate of Collagen I-Matrigel (Advanced BioMatrix, 5005) (ratio 4:1) with StemPro-34 SFM (Gibco, 10639011) supplemented with 15% FBS (Thermo, 10500064), 100ng/ml VEGFA and 100ng/ml FGF2 (Milteney Biotech, 130-093-841) to induce vascular network (VN) formation^6^. The sprouting appeared within two days in all preparations. Fresh media was provided after three days and then every other day. VNs were established between days 10 to 12 and extracted from the Matrigel to a 96-well ultra-low-attachment plate (2BScientific, MS-9096UZ) where they self-assembled to BVOs. BVOs were kept in culture for up to 40 days.

### Diabetic media treatment

For normoglycemia (‘Normal’) treatment, BVOs were cultured in StemPro-34 SFM Complete with 15% FBS, 100ng/ml VEGFA, and 100ng/ml FGF2 supplemented with 20mM D-Mannitol (AcrosOrganics, 125340010). D-Mannitol was used as an osmotic control^6^. A 7-day treatment was performed.

For diabetic media treatment, BVOs were cultured in StemPro-34 SFM Complete with 15% FBS, 100ng/ml VEGFA, and 100ng/ml FGF2 supplemented with 20mM D– (+)-Glucose (Sigma-Aldrich, G7021), 1ng/ml IL-6 and 1ng/ml TNFα^6^. The treatment started 3 days after VN extraction, and fresh media was provided every other day. As the media confer both hyperglycemia and low cytokine treatment they were termed ‘diabetic media’. A 7-day treatment was performed.

### iPS-derived Endothelial Cell (iPS-EC) and Mural Cell (iPS-MC) Differentiation

The method used was adapted from previously published protocols^11^. iPS cells were seeded on Corning Matrigel Growth Factor Reduced (GFR) Basement Membrane Matrix (SLS, 356231) at a density of 1.6×10^5^ cells per 6 well in StemMACS iPS-Brew XF (Milteney Biotech, 130-104-368) supplemented with 10 μM Rock inhibitor Y-27632 (ATCC ACS-3030) for 24 h. The following day (day 1), the medium was changed to N2B27 medium, a 1:1 ratio of Neurobasal medium (Thermo, 21103049) and DMEM/F12 (Gibco, 11330-032), with N2 (Thermo, 17502048), B27 (Thermo, 12587010), Glutamax (Thermo, 350050061) and freshly supplemented with 8 μM of CHIR (Sigma, SML1046) and 25 ng/ml of BMP4 (ThermoFisher, PHC9534). After 72h (day 4), the medium was replaced with StemPro-34 SFM (Gibco, 10639011) supplemented with 200 ng/ml human VEGF 165 (Peprotech, 100-20) and 2 μM Forskolin (R&D system, 1099). On day 6, cells were selected by Magnetic Activated Cell Sorting (MACS) for iPS-EC expressing CD144 using Microbeads Kit (Miltenyi, 130-097-857). Positive cells were seeded on mouse Collagen IV (Biotechne, 3410-010-02) coated plate in EGM2-MV medium (Promocell, C-22011) supplemented with 20% FBS (Thermo, 10500064) and 50 ng/ml VEGFA. Cells were used for experiments up to 3 passages. Negative CD144 cells (CD144^-^), hereafter referred to as iPS-derived mural cells (iPS-MCs), were cultured on 2% gelatin solution coated plates in EMB2 complete medium (Promocell, C-22011) supplemented with 2% FBS. Cells were used for experiments up to 3 passages.

### Seahorse Assays

Extracellular flux analysis studies were performed using a Seahorse XF^24^ analyzer (Agilent) according to the manufacturer’s protocol. Cells were plated on substrate-coated Seahorse XFe24 culture plates at 3 × 10^4^ cells/well with the respective substrate in complete media. For mitochondrial stress test, the media were changed 45 mins before the assay to unbuffered DMEM (pH 7.3) (Sigma-Aldrich, D5030) supplemented with either 10 mM glucose, 1 mM sodium pyruvate or 2mM glutamine, and the cells were incubated at 37 °C. Basal oxygen consumption rate (OCR), ATP-linked OCR, and maximal respiration were determined by the sequential administration of 1 µM oligomycin (Sigma, 75351), 2 µM carbonyl cyanide 4-

(trifluoromethoxy) phenylhydrazone (FCCP) (Sigma, C2920), and 1 µM antimycin A (Sigma, A8674) and 2 µM rotenone (Sigma, R8875). For fatty acid oxidation (FAO) experiments, 1 mM BSA-conjugated palmitate was added in each well just before starting the experiment.

For glycolysis, the media was changed 45 mins before the assay to unbuffered DMEM (Sigma-Aldrich, D5030) pH 7.4, at 37 °C, 1 h before assay and the extracellular acidification rate (ECAR) was determined at baseline, and after the sequential injection of 10 mM glucose, 1 µM oligomycin (Sigma, 75351), and 50 mM D-deoxy D-Glucose (Sigma, D6134). At the end of the experiment cells were fixed using 4% paraformaldehyde (PFA) for 15 min and treated with 5 µM DRAQ5^TM^ (Thermo Scientific, 62251) for 20 min. Cell nuclei were imaged and analysed using Li-cor Odyssey imager and quantification was used as an index of cell number for data normalization.

### Immunohistochemistry

BVOs were rinsed twice in PBS and then fixed in 4% PFA for 1 h at room temperature (RT) and washed twice in PBS. BVOs were dehydrated in 20% sucrose at 4°C overnight, then embedded in gelatin solution and stored at –20°C for cryosectioning. The frozen BVOs were sectioned using NX70 Cryostat (Thermo Scientific) to 20 thickness. Frozen sections were then washed in PBS for 5 minutes and then permeabilized and blocked in Blocking buffer for 2h at RT. Primary antibodies were diluted in blocking buffer and incubated overnight in a cold room. Antibodies used and dilution factors’ information are available in Supplementary Table 3. On the following day, sections were washed three times in PBS with 0.1% Tween 20 (PBS-T) and then incubated with labelled secondary antibodies for 2 h at RT. After two washes in PBS-T the sections were counterstained with DAPI and mounted with Fluoromount-G mounting media (Thermo, 00-4958-02).

Vessel density and length were quantified using Vessel Analysis Plugin on ImageJ (Fiji, version 2.0.0) as a percentage (%) of the area analysed. ColIV deposition was quantified using ImageJ (Fiji, version 2.0.0). Stack images were resliced along a vertical axis and cross-sections were used for quantification.

### Quantitative Reverse Transcription PCR (qRT-PCR)

RNA was isolated using the PureLink RNA Mini kit (ThermoFisher Scientific, 12183018A) according to the manufacturer’s recommendation. RNA was reverse-transcribed into cDNA using High-Capacity cDNA Reverse Transcription Kit (ThermoFisher, 4368813) and qRT-PCR reactions were performed using Taqman gene expression assays and Taqman Mastermix (Thermo Fisher Scientific, 4444557) on a StepOnePlus Real-Time PCR System with the following cycling parameters: 95°C for 10 minutes, 40 cycles of 95°C for 15 seconds and 60°C for 1 minute. Beta-Tubulin was used as a normalisation control.

### Flow Cytometric Analysis

BVOs were mechanically dissociated using a scalpel and then incubated in Dissociation solution (1.7mg Dispase, 0.2mg Liberase and 0.1mg DNase per ml) in PBS for 20 min at 37°C. BVO solutions were passed up to 10 times through 21g needles. Approximately 50,000 single cells were resuspended in 100μl of FACS buffer (PBS containing 1% FBS) and stained with fluorescence-conjugated antibodies for 30 min at 4°C in the dark. The antibodies used and dilution factors were according to a previous publication^5^. The cells were washed in PBS and resuspended in 1% PFA in PBS. Data were acquired the following day using a Fortessa Flow Cytometer analyzer (BD) and analyzed using FlowJo software (Becton & Dickinson and Company). Antibodies used and dilution factors’ information are available in Supplementary Table 4.

### Genome editing using CRISPR-Cas9

A single guide RNA targeting the human IGFBP7 was designed using the CRISPR Finder online tool (Wellcome Sanger Institute Genome Editing) and cloned into a mammalian expression vector encoding Cas9 from *S. pyogenes* and a puromycin resistance gene (Addgene, 62988) using Golden gate reaction^5^. The following primers were used for IGFBP7 guide RNA cloning:

IGFBP7 F: CACCGGACGGCACCACCTACCCGAG

IGFBP7 R: AAACCTCGGGTAGGTGGTGCCGTCC

The sgRNA vector was confirmed by Sanger sequencing and transfected into iPS cells using Lipofectamine 3000. In brief, 5 µg of plasmid were transfected into iPS cells seeded on matrigel-coated six-well plates in StemMACS iPS Brew-XF media (Miltenyi Biotech, 130-104-368) supplemented with 10 μM Y-27632 for 24h. The following day, the transfection complexes were removed and fresh StemMACS iPS Brew-XF media supplemented with 10 μM Y-27632 was added to the cells. Puromycin selection (0.2 µg/ml) was performed for 48h, and single cells were cultured for an additional ten days. Single colonies representing isogenic mutant lines were selected and expanded. Knockout cell lines were verified by Sanger sequencing and ELISA assay.

### Proteomic Analysis of the BVOs

Proteomic analysis was performed at the King’s Smart Trials Hub Facility. For all proteomic experiments, the KOLF2 iPS line was used to generate BVOs.

The protein samples were thawed on ice and diluted on an average 4-fold in 50 mM triethylammonium bicarbonate (TEAB). Nineteen micrograms of proteins from each sample were withdrawn according to the protein concentration shared and made up to 60 µL with 50 mM TEAB. The disulphide bridges of the proteins in sample were reduced with 4 mM dithiothreitol (DTT) by incubating at 56°C for 10 min shaking continuously. The reduced proteins were alkylated with 14 mM Iodoacetamide (IAA) by incubating in dark for 30 min to avoid reannealing of the reduced disulphide bridges. Excess IAA is quenched by raising the DTT concentration to 7 mM. The proteins were digested overnight at 37°C at 1:100 enzyme: protein ratio. After the digestion the trypsin activity was quenched by the addition of 1% trifluoracetic acid (TFA) and the surfactant was hydrolysed by incubating at 37°C for 45 min. The hydrolysed RapiGest along with impurities, if any, were removed by centrifugation at 12,000 g for 15 min at 4°C. The harvested peptides were dried under speed vac. The dried peptides were resuspended in 0.1% TFA containing 2% Acetonitrile (ACN) by bath sonication.

A preliminary survey run with the digested peptides was carried out to determine the on-column loading concentration. As informed by the survey run, 400 ng of peptides from each sample were analysed on Orbitrap Astral mass spectrometer (Thermo Fischer Scientific) coupled in line with a Vanquish Neo nanoLC (Thermo Fischer Scientific) chromatography system employing a nDIA approach. The peptides were loaded onto a trapping column (pepmap neo, 0.5 cm x 300 µm, Thermo Fisher Scientific), washed to remove contaminants, if any, and separated on a 50 cm long C18 nano column (EasySpray, 75 µm ID, 1.9 µm particle size, 120 Å pore size, Thermo Fisher Scientific) at 300 nL/min flow rate using a gradient method. 0.1% formic acid (FA) in water was used as mobile phase A and 0.1% FA in 80% acetonitrile (ACN) was used as mobile phase B to create the gradient. The column was incubated at 45°C throughout the run. The eluted peptides were ionized at 1.9 kV spray voltage, and the precursor peptide mass spectra (MS1) were acquired on an Orbitrap analyzer at 120,000 resolving power across the mass range 380-980 Th. The fragmentation spectra (MS2) were acquired in parallel on the Astral analyzer after fragmenting all the precursors selected at 3Th isolation window by the HCD method. The fragmentation spectra from the whole mass range were acquired across 200 scan events. The normalized collision energy used for the HCD fragmentation was 32%. The ions for each MS2 fragmentation spectra were accumulated over 5ms. Each DIA run was 60 min long. At the end of each run, the columns were washed with 3-column volumes of 95% mobile phase B and equilibrated with 3-column volumes of 5% mobile phase A. The samples from each set were acquired in a randomized order and blanks were included after every 3 runs to avoid carryover effects, if any. The system suitability was assessed by injecting 50 ng HeLa digest QC standard.

### Proteomic Analysis of the BVO secretome

Secreted proteins were concentrated using 3kD MWCO spin filters (Amicon, Merck, #10212584) at 20,000 x g, 4°C. The protein samples were denatured by 6M urea, 2M thiourea and reduced by 10mM DTT at 37°C for 1h, alkylated by 50mM iodoacetamide at RT in the dark for 1h, and washed with 0.1M TEAB, pH=8.5 for 3 times in the spin filter. The proteins were digested by trypsin/lysC (protein: enzyme = 25:1, Promega, #V5071) at 37°C overnight. Peptides were acidified by 0.1% TFA and C18 cleanup was performed using C18 cartridges (Thermo Fisher Scientific, #89870) following the manufacturer’s instruction. Eluted peptides were dried to completion in a SpeedVac (Thermo Fisher Scientific) and resuspended in 2% Optima ACN, 0.05% Optima TFA in LC-MS grade H_2_O (Fisher Scientific).

Samples were separated by an UltiMate3000 nanoRSLC system. Peptides were loaded onto a trapping column (Pepmap Neo, 0.5 cm x 300 µm, Thermo Fisher Scientific, #174500) and separated on a 50cm column (EASY-Spray C18 reversed-phase column, 75µm x 50cm, 2 µm, Thermo Fisher Scientific, ES75500PN) at 250 nl/min using the following LC gradient: 0-10 min: 4% B; 10-75 min: 4-30 B; 75-80 min: 30-40% B; 80-85 min: 40-99% B; 85-89.8 min: 99% B; 89.8-90min: 99-4% B; 90-120 min: 4% B (A=0.1% formic acid in H_2_O, B=80% ACN, 0.1% formic acid in H_2_O). The separated peptides were directly injected into an Orbitrap Eclipse Mass Spectrometer (Thermo Fisher Scientific) and ionised with 1.9 kV spray voltage and ion transfer tube temperature set at 275°C. Full MS spectra were collected using Orbitrap with scan range of 350-1600 m/z, resolution of 120,000, AGC 4.0e5 and maximum injection time set to 50 msecs. The 20 most abundant ions from the full MS scan were selected for data-dependant MS2 with HCD fragmentation and acquired using the Orbitrap analyser with a resolution of 60,000, AGC 5.0e4, normalised collision energy set to 38%, isolation windows 0.7 m/z and maximum injection time set to 105 msecs. Dynamic exclusion of 35 seconds and lock mass of 445.12003 m/z were used. A blank was analysed after each sample, and the system performance was assessed using 50ng HeLa digest standard (Thermo Fisher Scientific, #88328).

### Proteomics Data Processing and Protein Quantification

Raw DIA data from the diabetic BVO and secretome datasets were analysed in library-free mode using Proteome Discoverer (version 3.1; Thermo Fisher Scientific) with the CHIMERYS algorithm (version 2.0; MSAID GmbH), MaxQuant (version 2.7.5) with the predicted library mode in the MaxDIA workflow, FragPipe (version 23.1) with the MSFragger-DIA workflow (version 4.3) incorporating DIA-NN (version 1.8.2 Beta 8) for quantification. Searches were performed against the UniProt/SwissProt human and bovine protein database (version 2025_04; 26,472 proteins). Trypsin was specified as the proteolytic enzyme with a maximum of two missed cleavages. Carbamidomethylation of cysteine was set as a fixed modification, while methionine oxidation was included as a variable modification. For CHIMERYS, fragment ion mass tolerance was set to 20 ppm. For MaxQuant, first-search and main-search peptide mass tolerances were set to 20 ppm and 4.5 ppm, respectively. For MSFragger-DIA, precursor and fragment mass tolerances were set to 10 ppm and 20 ppm, respectively, with mass calibration and parameter optimisation enabled. Identifications were filtered at a 1% false discovery rate (FDR) at all levels.

Raw DDA data from the PFKFB3 knockout BVO secretome dataset were reprocessed using Proteome Discoverer (version 3.1; Thermo Fisher Scientific) with the SEQUEST-HT algorithm, MaxQuant (version 2.7.5) with the Andromeda search engine, and FragPipe (version 24.0) with the MSFragger algorithm (version 4.4). Searches were performed against the UniProt/SwissProt human and bovine protein database (version 2021_01; 26,431 proteins). Trypsin was specified as the proteolytic enzyme with a maximum of two missed cleavages. Carbamidomethylation of cysteine was set as a fixed modification, while methionine oxidation was included as a variable modification. For SEQUEST-HT and MSFragger, precursor and fragment mass tolerances were set to 10 ppm and 20 ppm, respectively. For MaxQuant, first-search and main-search peptide mass tolerances were set to 20 ppm and 4.5 ppm, respectively, with match-between-runs enabled. Identifications were filtered at a 1% FDR at all levels.

Raw DIA data from the IGFBP7 knockout BVO dataset were analysed in library-free mode using Spectronaut (version 20.3; Biognosys) with the directDIA+ (deep) mode in the directDIA workflow, MaxQuant (version 2.7.5) with the predicted library mode in the MaxDIA workflow, FragPipe (version 24.0) with the MSFragger-DIA workflow (version 4.4) incorporating DIA-NN (version 1.8.2 Beta 8) for quantification. Searches were performed against the UniProt/SwissProt human protein database (version 2025_04; 20,659 proteins). Trypsin was specified as the proteolytic enzyme with a maximum of two missed cleavages. Carbamidomethylation of cysteine was set as a fixed modification, while methionine oxidation and protein N-terminal acetylation were included as variable modifications. For MaxQuant, first-search and main-search peptide mass tolerances were set to 20 ppm and 4.5 ppm, respectively. For MSFragger-DIA, precursor and fragment mass tolerances were set to 10 ppm and 20 ppm, respectively, with mass calibration and parameter optimisation enabled. Identifications were filtered at a 1% FDR at all levels.

All quantitative outputs were normalised using the built-in methods implemented within each platform and exported for downstream analyses.

### Bioinformatic analysis

Biological replicates were performed in all the experiments with n = 3 independent preparations. Data are expressed as the mean ± SD and were analysed using GraphPad Prism 9 software with a two-tailed Student’s t-test for two groups or ANOVA for more than two groups. A value of *p < 0.05, **p < 0.01, ***p < 0.001, ****p < 0.001 was considered significant.

For proteomics, datasets analysed using combined human and bovine protein databases, proteins with preferential peptide-spectrum match support for bovine homologues were excluded. Datasets were subsequently filtered to retain proteins with less than 30% missing values. Remaining missing values were imputed using KNN-impute, with k set to the number of samples in the smaller phenotype group minus one. Protein abundances were log2-transformed prior to downstream analyses.

Differential expression (DE) analyses of mass spectrometry proteomics data were performed using the limma package^46^ with eBayes moderation and adjustment for selected covariates. For each dataset, DE outputs from the platform yielding the highest number of protein identifications were combined with non-overlapping proteins from other platform outputs demonstrating adequate agreement (Spearman’s rho > 0.4 and DE directional concordance > 0.7) to improve proteome coverage. Nominal p-values were corrected using the Benjamini–Hochberg method^47^. Due to the limited sample size, significance was defined as nominal p < 0.05 and absolute log2 fold change > 0.3.

Protein regulatory network reconstruction was performed using the directional regulatory network reconstruction with adaptive partitioning (DiRec-AP) pipeline with 100 bootstrap iterations, a method that combined Conditional Mutual Information with adaptive partitioning (Aracne-AP)^48^ to reconstruct protein regulatory network with the SIREN tool^49^ tool to classify connections into activation or inhibition ones (available at https://github.com/Cardiovascular-Bioinformatics/Direc-AP). Significant directional interactions were defined using a Benjamini–Hochberg adjusted p-value threshold of 0.05. The directional network reconstructed from the diabetic BVO secretome dataset was subsequently integrated with the network reconstructed from the PFKFB3 knockout BVO secretome dataset to enhance network coverage and regulatory connectivity, with interaction directionality retained from the diabetic BVO network in cases of conflict.

Transcription factor (TF) activities were inferred from proteomics secretome data of BVO using DoRothEA regulons (confidence levels A–B) and the VIPER algorithm implemented in the decoupleR R package^50–52^. VIPER estimates TF regulatory activity by assessing coordinated expression changes of curated target genes, providing a quantitative activity score per TF and sample. TF activity differences between IGFBP7 knockout and control BVO samples were assessed using unpaired two-sided Student’s t-tests applied to VIPER-derived activity scores for each TF. P-values were adjusted for multiple testing using the Benjamini–Hochberg procedure, and TFs were ranked by adjusted significance. Effect sizes were calculated as the difference in mean activity between conditions. Significantly altered TFs (FDR < 0.05) were visualized as a row-scaled heatmap of VIPER activity scores. Hierarchical clustering was applied to TFs, while samples were ordered by condition without clustering to preserve experimental structure.

Gene set enrichment analysis was performed in R using the fgsea package (v1.28.0) on ranked differentially expressed proteins identified by mass spectrometry in IGFBP7 knockout versus control BVO samples. Hallmark gene sets from the Molecular Signatures Database (MSigDB) were used, and pathways were ranked according to normalized enrichment score (NES).

Volcano plots and enrichment dot plots were generated using the ggplot2^53^ package within the R programming environment^54^ (version 4.5.2). Network visualisations were performed using Cytoscape^55^. Pathway and functional enrichment analyses were conducted using DAVID^56^, including pathway annotations from Reactome^57^ and KEGG^58^, as well as functional annotations from Gene Ontology^59^. Significantly enriched terms were defined using a Benjamini–Hochberg adjusted p-value threshold of 0.05.

Time-to-event associations with incident cardiovascular disease (CVD) and heart failure (HF) were assessed using Cox proportional hazards (CoxPH) regression. Diabetes, CVD and HF status and endpoint definitions were obtained from Oexner et al. (2025)^60^. Hazard ratios (HRs) and 95% confidence intervals (CIs) were estimated for each predictor, and model assumptions were assessed where appropriate.

Nephropathy / Kidney Disease status was inferred from the UK Biobank by checking the ICD-10 codes: Glomerular disorders in diseases classified elsewhere (N08), Chronic kidney disease (CKD) (N18), Unspecified kidney failure (N19), Diabetes with kidney complications (diabetic nephropathy) (E10.2 / E11.2) and the ICD-9 codes: CKD (585), Renal failure, unspecified (586). Retinopathy status was inferred by checking the ICD-10 codes: Diabetic retinopathy (H36.0) and Other retinal disorders (H35.0-H35.9) and the ICD-9 code diabetic retinopathy / macular edema (362.0x). ICD codes were also used to classify patients into T1D, T2D and non-diabetics, with T1D patients being the ones with ICD-10 diagnosis of Insulin-dependent diabetes mellitus(T1D) (E10.0-E10.9) or ICD-9 diagnosis of Diabetes mellitus, type I (IDDM) manifestations (250.x1 and 250.x3) and T2D being the ones with ICD-10 diagnosis of Type 2 diabetes mellitus (E11.0-E11.9) or ICD-9 diagnosis of Type II diabetes mellitus (250.x0, 250.x2). T1D and T2D status were ascertained at baseline using UK Biobank (UKB) data; however, the timing of disease onset before baseline recruitment was unavailable, precluding estimation of diabetes duration.

Genetic variants located within ±500 base pairs of the genomic regions of the IGFBP7, CD93, LGALS1, and LGALS3 genes were identified and functionally annotated using the Ensembl Variant Effect Predictor (VEP). Variants were selected based on their proximity to the corresponding gene loci, including potential regulatory regions adjacent to the transcribed sequence. To assess potential clinical relevance, annotated variants were cross-referenced with the ClinVar database, and variants classified as clinically significant or previously associated with disease phenotypes were retained for downstream analyses. To further investigate potential regulatory effects on gene expression, variants were interrogated against expression quantitative trait loci (eQTL) datasets from the Genotype-Tissue Expression (GTEx) Project. Variants demonstrating significant eQTL associations (with nominal p-values less than 0.05) in tissues relevant to cardiovascular and renal biology, including kidney, heart, whole blood, and blood vessel tissues, were identified and prioritized. The genetic variants were further analysed using whole genome sequencing data from the All of Us cohort. The variants were associated with prevalent and ever diagnosed diabetes disease (T1D and T2D) and diabetic microangiopathy complications (retinopathy and nephropathy) as well as with future 5-year cardiometabolic outcomes (diabetes, CVD and HF) in a CVD healthy at baseline subpopulation. Endpoints were defined with the same combination of ICD-9 and 10 codes as in UKB while both odds ratio and hazard ratio analysis were corrected for Age and Sex effects.

## Notes

### Competing Interest Statement

The authors have declared no competing interest.

### Author Declarations

UK Biobank; All of Us; The iPS lines generated from peripheral blood mononuclear cells of healthy donor (non-diabetic) (HD) and diabetic (DB) donors as described previously were also used (ethical approval from the Office for Research Ethics Committees of Northern Ireland (REC 14/NI/1109)).

